# PASHiOn: Personalised Against Standard High tibial Osteotomy, a prospective multi-centre randomised controlled trial: Trial Protocol

**DOI:** 10.1101/2025.06.29.25330037

**Authors:** Harinderjit Singh Gill, Alisdair MacLeod, Lisa Poulton, David Smith, Heidi Fletcher, Vipul Mandalia, Andrew Toms, Patrick Hourigan, James Murray, Allister Trezies, Chris Wilson, Loretta Davies, David Beard, Nicholas Peckham, Ines Rombach, Jonathan Cook

## Abstract

Knee osteoarthritis (OA) is common, painful, progressive and disabling, and has significant personal and societal burden, particularly in an ageing population. Whilst knee replacement is successful for very late stage disease, it is inappropriate for earlier stages, leaving few effective treatments available for patients in the “treatment gap” between symptom free and late-stage arthritis. High Tibial Osteotomy (HTO) is a proven surgical treatment providing good long-term outcome, reducing pain and improving function. HTO involves changing the alignment of the tibia to correct improper joint loading. Outcomes of HTO surgery are linked directly to the accuracy of the surgical re-alignment with under-correction resulting in worse outcomes. Inaccuracies occur primarily due to limitations of the current planning and surgical technique. A new method (TOKA) has been devised involving personalisation and digital planning. This method uses a custom personalised surgical guide and plate, tailored to the patient.

The aim of the PASHiOn trial is to establish whether digitally planned personalised HTO surgery (TOKA) increases the accuracy of bone correction in comparison to conventional HTO surgery. Embedded within the trial is a nonrandomised pre-RCT technology check and safety assessment of 5 patients (Phase 1), followed by a randomised controlled trial of 88 patients (Phase 2).

During Phase 1 of the clinical investigation, 5 patients fulfilling the inclusion criteria will be recruited and assessed in an identical way to the 88 patients recruited in the main trial, but without randomisation. Recruitment of the remaining patients (Phase 2) will take place after the six-week assessment on the fifth patient is complete and the oversight committee supports progression to Phase 2. Patients in Phase 1 and 2 will be followed up in the same manner for a period of up to 15 months from registration (Phase 1) and randomisation (Phase 2) (12 months from surgery).

## BACKGROUND AND RATIONALE

A list of abbreviations used is provided as Table 1 and the synopsis for the trial is given in Table 2. Osteoarthritis (OA) of the knee carries a huge personal and societal burden. Up to 28% of the UK population over 40 have knee pain, with half of these people having radiographic OA (1). This high prevalence of disease is reflected in the demand for knee replacement (>108,000/year in UK). It is also important to recognise that a third of knee replacements are now performed in patients younger than 65 (2) indicating the disease burden and need for treatment in a younger group of OA sufferers. Hermans et al (3) estimated the annual cost of conservatively treated symptomatic knee OA to be £9,300 per individual; the majority (80%) of this cost is due to loss in productivity, the remainder due to medication. Within the 40 to 65 years age group (21 million people), 2.9 million people of working age have radiographic knee OA resulting in annual burden of over £26 billion. With population ageing (4), the UK demand for substantive intervention such as HTO and knee replacement is set to more than double by 2030 (5).

**Table 1.** List of Abbreviations.

|  |  |
| --- | --- |
| **ADE** | **Adverse Device Effect** |
| **AE** | **Adverse Event** |
| **AR** | **Adverse Reaction** |
| **BMI** | **Body Mass Index** |
| **CA** | **Competent Authority** |
| **CAD** | **Computer-aided design** |
| **CI** | **Chief Investigator** |
| **CIP** | **Clinical Investigation Plan** |
| **CRF** | **Case Report Form** |
| **CT** | **Clinical Trials** |
| **CTA** | **Clinical Trials Authorisation** |
| **DD** | **Device Deficiency** |
| **DSMC** | **Data and Safety Monitoring Committee** |
| **DSUR** | **Development Safety Update Report** |
| **eCRF** | **Electronic Case Report Form** |
| **GCP** | **Good Clinical Practice** |
| **GP** | **General Practitioner** |
| **HKA** | **Hip Knee Ankle** |
| **IR(ME)ER** | **Ionising Radiation Medical Exposure Regulations** |
| **HRA** | **Health Research Authority** |
| **HTO** | **High Tibial Osteotomy** |
| **IB** | **Investigators Brochure** |
| **ICF** | **Informed Consent Form** |
| **ICH** | **International Conference on Harmonisation** |
| **IMD** | **Investigational Medical Device** |
| **IP** | **Intellectual Property** |
| **ISF** | **Investigator Site File** |
| **KOOS** | **Knee injury and Osteoarthritis Outcome Score** |
| **KOS** | **Knee Outcome Survey** |
| **MHRA** | **Medicines and Healthcare products Regulatory Agency** |
| **MSKBRF** | **Musculoskeletal Biomechanics Research Facility** |
| **NHS** | **National Health Service** |
| **NIDJD** | **Non-Inflammatory Degenerative Joint Disease** |
| **NIHR** | **National Institute for Health Research** |
| **NRES** | **National Research Ethics Service** |
| **NYHA** | **New York Heart Association** |
| **OA** | **Osteoarthritis** |
| **OCTRU** | **Oxford Clinical Trials Research Unit** |
| **OKS** | **Oxford Knee Score** |
| **PACS** | **Pain Audit Collection System** |
| **PI** | **Principal Investigator** |
| **PIS** | **Participant/ Patient Information Sheet** |
| **PPI** | **Patient and Public Involvement** |
| **PROMs** | **Patient Reported Outcome Measures** |
| **QA** | **Quality Assurance** |
| **R&D** | **NHS Trust R&D Department** |
| **RCT** | **Randomised Controlled Trial** |
| **REC** | **Research Ethics Committee** |
| **ROM** | **Range of Motion** |
| **RRAMP** | **Registration/Randomisation and Management of Product** |
| **SADE** | **Serious Adverse Device Effect** |
| **SAE** | **Serious Adverse Event** |
| **SAP** | **Statistical Analysis Plan** |
| **SDV** | **Source Data Verification** |
| **SITU** | **Surgical Intervention Trials Unit** |
| **SOP** | **Standard Operating Procedure** |
| **SUSAR** | **Suspected Unexpected Serious Adverse Reactions** |
| **TMF** | **Trial Master File** |
| **TOKA** | **Tailored Osteotomy Knee Alignment** |
| **TSC** | **Trial Steering Committee** |
| **TSI** | **Trial Specific Instruction** |
| **UCLA Activity Score** | **University of Carolina, Los Angeles Activity Score** |
| **UK CRN** | **UK Clinical Research Network** |
| **UKR** | **Unicompartmental Knee Replacement** |
| **USADE** | **Unexpected Serious Adverse Device Effect** |
| **USM** | **Urgent Safety Measure** |
| **VAS** | **Visual Analogue Scale** |
| **VVA** | **Video Vector Analysis** |
| **3D** | **Three Dimensional** |

**Table 2:** Trial Synopsis.

|  |  |
| --- | --- |
| **Study Title** | **PASHiOn: Personalised Against Standard High tibial Osteotomy, a prospective multi-centre randomised controlled trial** |
| **Type of study** | **Pre market** |
| **Trial Design** | **Multi-centre, blinded, superiority two arm, parallel group design randomised controlled trial** |
| **Trial Participants** | **Patients undergoing High Tibial Osteotomy (HTO)** |
| **Planned Sample Size** | 93 patients, revised to 55<br>Phase 1: Initial safety phase: 5 patients<br>Phase 2: RCT: 88 patients, revised to 50 patients |
| **Follow-up duration** | 9 months after randomisation, (minimum 4 months after surgery). (Some secondary outcomes (PROMS) 15 months from randomisation*) |
| **Planned Trial Period** | 42 Months. Extended to 66 months. |
| **Primary Objective**<br>**(Phase 1)** | To assess the safety of the TOKA device. |
| **Primary**<br>**Endpoint**<br>**(Phase 1)** | Incidence/rate of safety events – AEs, ADEs, SAEs, SADEs, USADEs and complications. |
| **Primary Objective**<br>**(Phase 2)** | To establish if digitally planned personalised HTO surgery (TOKA) increases the accuracy of bone correction in comparison to conventional HTO surgery. |
| **Primary Endpoint**<br>**(for analysis)** | The absolute difference between planned and achieved coronal plane correction in terms of the hip-knee ankle angle, at nine months post-randomisation (six months post-surgery) as measured by full weight bearing X-rays. |
| **Secondary**<br>**Objective** | To compare the treatment arms in terms of: |
| **Secondary**<br>**Endpoints** | Number of patients (%) achieving a pre-specified difference between planned and achieved coronal plane correction at six months post-surgery; change in coronal plane location of peak loading during gait measured by video vector comparing 9 month (post randomisation) relative to baseline; operative time in minutes; Range of Motion; patient reported scores at 9 (all) and 15 months (funding allowing) : KOOS score, generic health related quality of life (EQ-5D-5L), VAS pain score, satisfaction and transition, health resource use. Relationship between correction angle, change in loading calculated by computer modelling and outcome. |
| **Device Name** | Tailored Osteotomy Knee Alignment (TOKA<sup>®</sup>) |
| **Manufacturer**<br>**Name** | 3D Metal Printing Ltd, The Innovation Centre, University of Bath, BA1 1UB |
| **Principle intended**<br>**use** | The product is a customised medical device for use in opening wedge high tibial osteotomy (HTO). |
| **Length of time use**<br>**the device has been**<br>**in use.** | n/a – although TOKA is a new device, it is substantially similar to existing devices and intended for use during a well-established procedure |
|  | The comparators, Tomofix and ActivMotion are the most widely used devices for HTO procedures in the UK. Surgical procedures using the Tomofix and ActiveMotion are similar to TOKA, however, they are not patient specific and do not use custom-made plates. |

The current treatment of choice for late stage OA is total knee replacement, however, it is not recommended for the earlier stages of knee OA and it does not last as long in young patients. Patients aged 64 years or less at time of primary surgery have double the revision rate of those aged between 65 and 74 years (6). The stage or grade of OA, mainly based on radiographs (x-rays) (7), is a key factor used by surgeons when deciding whether to perform knee replacement (8). There is a poor relationship between symptoms and radiographic grade (9). A significant number of individuals find themselves in the treatment gap, suffering pain and disability from knee OA but are not yet suitable for knee replacement (10).

An alternative to knee replacement is high tibial osteotomy, commonly referred to as HTO. This procedure is suitable for isolated medial compartment knee OA which occurs in up to 29.5% of all knee OA cases (11, 12). In HTO, the native joint is preserved by re-aligning the tibia to off-load the worn areas of the knee. An opening wedge is made in the tibia which is then stabilised by a metal plate. A large body of evidence demonstrates that HTO performed with correct indications and surgical technique has good long-term results (13, 14). Well-performed HTO can delay the need for knee replacement by 10 years and in a considerable number of cases it can be the definitive treatment, thereby avoiding the need for further surgery altogether (15–17). Motion analysis studies have demonstrated that HTO normalises walking patterns and significantly reduces knee adduction movement at six months post-surgery (18–20). Recent work by Besselink et al (21) has shown that medial opening wedge HTO leads to a statistically significant increase in the medial joint space width (0.52 mm) at one-year post-surgery compared to pre-operative values. At two years, the medial joint space width increased to 1.05 mm. This study indicates that some recovery of the cartilage can occur after HTO.

There are a number of barriers to wider adoption of HTO. The key surgical barrier is the challenging operative technique, which carries a substantial risk of not being able to achieve the planned bone correction and hence the desired change in the location of the knee loading. The gold standard for assessing lower limb alignment in the coronal plane is to measure the hip-knee-ankle angle on full-length weight bearing x-rays (22). Van Den Bempt et al performed a systematic review of the accuracy of coronal plane correction (23). They proposed an optimal accepted accuracy range of between 2 to 6 degrees of valgus for the coronal limb alignment correction after HTO yet reported only about 50% of conventional HTO operations achieve this optimal range of alignment correction. When correction was not within the optimal range, there was a tendency to under-correct. El-Azab et al have shown that under-correction is associated with significantly worse clinical outcomes than accurate correction and overcorrection (24). Van Den Bempt et al make two key conclusions: 1. “the accuracy of alignment corrections for conventional HTO methods is poor” and 2. “Although HTO procedures have been shown to be very successful in the treatment of unicompartmental knee arthritis when performed accurately, the results of this review stress the importance of ongoing efforts in order to improve correction accuracy in modern HTO”. The results of the operation when not performed accurately are far less satisfactory.

A new HTO procedure called (ToKa: Tailored osteotomy for Knee alignment) has been developed with funding from an Arthritis Research UK (now Versus Arthritis) Medical Technologies grant. The ToKa procedure is tailored to each patient and the mechanical safety has been demonstrated using a combination of experimental testing and computer simulation (ToKa virtual or in silico trial).

With the ToKa technology, three-dimensional (3D) CT-based digital planning leads to the auto-generation of a personalised contoured plate and a personalised surgical guide, which are then 3D printed in medical grade titanium alloy. The 3D printing process is highly accurate, better than 0.1 mm. All the screw orientations and lengths are determined during the 3D planning and incorporated into the personalised surgical guide, removing the need for in situ measurements. The personalised surgical guide fits to the patient’s bone and has a slot defining the location and orientation of the tibial cut. The guide removes the need for inter-operative radiology and reduces the surgical time needed. The key driving factor in the design of the ToKa procedure, with its personalised guide and plate, was to achieve high accuracy in creating the planned correction. The difference between ToKa and generic HTO is akin to the difference between a made to measure suit and an “off the peg” suit; the “off the peg” suit can be an acceptable fit, but never quite matches that of the tailor-made version. Generic HTO plates do not conform to the tibial surface and result in skin stretching over the plate, giving rise to soft tissue irritation. A major reason for patient dissatisfaction is due to soft tissue irritation; studies have reported 40% of generic plates are removed for local soft tissue irritation (25, 26).

Based on our literature review, previous research funded by Arthritis Research UK (now Versus Arthritis), and in response to feedback from our Patient Group, a study to compare the accuracy of achieving the planned correction between the ToKa HTO procedure and standard generic HTO procedure is needed to see if this new procedure will lead to greater accuracy and repeatability of lower limb alignment correction. A repeatable accurate HTO procedure will make this treatment a viable option for patients with early to mid-stage knee OA. This study also has the potential to demonstrate that HTO surgery can be performed in a straightforward simple manner and would thus enable more patients to be offered the surgery.

A repeatable procedure that can achieve the planned correction also offers the potential to optimise the HTO procedure. Whilst HTO is effective when under-correction is avoided, the exact required amount of correction remains unknown. Having a cohort of patients for whom CT data, video vector analysis and clinical outcomes were available would allow computer models to be created in order to calculate the change in loading achieved for a given correction and relationship with outcome to be examined.

## IDENTIFICATION AND DESCRIPTION OF THE INVESTIGATIONAL DEVICE

### The Investigational Device and Intended Purpose

The investigational device is: Tailored Osteotomy Knee Alignment (TOKA^®^) version 1.0

The Manufacturer is: 3D Metal Printing Ltd, The Innovation Centre, University of Bath, BA1 1UB, UK

The device is registered with MHRA as: Device ID number: 2019011801169017

The device is a class IIb according to Medical Device Directive 93/42/EEC and MEDDEV 2. 4/1 Rev. 9 (MDD) Annex IX classification:

*“Rule 8: All implantable devices and long-term surgically invasive devices are classified as Class IIb.*

The Global Medical Device Nomenclature (GMDN) code to describe and catalogue TOKA® is 61689 (Internal orthopaedic fixation system, plate/screw, non-bioabsorbable, non-sterile)

The investigational device, TOKA, is intended to be used for the treatment of medial-compartmental tibiofemoral osteoarthritis. The device is a digitally planned (using individual CT scan measured anatomic data) personalised opening wedge high tibial osteotomy (HTO) procedure using a custom 3D printed surgical guide and plate. For the personalised treatment, surgical planning determines the correction angle and this, in combination with the 3D geometry obtained from the CT scan, is used to create the specific geometry of the surgical guide and conforming plate, which is then 3D printed in the titanium alloy metal.

3D Metal Printing Ltd manufacturing operations and facilities follow EN ISO 13485 and the Quality System Regulations (21 CFR 820) for design, development, manufacture, and distribution of medical devices. Components are outsourced with custom or off-the-shelf specifications and manufactured by qualified outside suppliers. A detailed overview of the manufacturing process is outlined in the TOKA Investigators Brochure.

### Device Traceability

Every TOKA device is given a uniquely identifiable, but anonymised, case number based on the date the CT was received from the hospital. All lot/batch numbers from components used in the TOKA set are recorded individually by the manufacturer (3D Metal Printing Ltd) on the device history file (DHF) in accordance with their ISO 13485 quality management system.

Product Labels include:

- Address of Manufacturer
- Handling Instructions
- Any special Instructions, including ‘Non-sterile’ if sterilisation is to be performed by the customer
- ‘Exclusively for clinical investigation’ if the product is intended for Clinical investigations of medical devices
- ‘Custom-Made Medical Device’
- PASHiOn Patient ID number
- Product ID number
- Use by date and date of manufacture

Component labels will contain the ID number only.

Full traceability is assured through issuing appropriate documentation and maintaining records that accompany every step-in batch manufacturing. Records of the CAD design work and all key variables used are kept. All data is maintained by 3D Metal Printing for a minimum of 15 years from receipt according to the ISO 13485 Quality Management System implemented by 3D Metal Printing Ltd.

### Description of the Investigation Device

TOKA® is indicated for patients with:

- Uni-compartmental medial osteoarthritis of the knee
- Varus deformity less than 20°

Contraindications include:

- BMIl1over 35
- Known deep tissue sensitivity to device materials

Materials used in the TOKA device that may come into contact with the body are described in the TOKA Investigator brochure. The TOKA^®^ System is not manufactured from materials/substances derived from animal or human origin.

#### User Training and Experience

Training of all surgeons in the TOKA procedure will take place prior to recruitment, at the Vesalius Clinical Training Unit at the University of Bristol, where full cadaveric facilities are available. The training will be organised by the CI and led by the PI at the lead site, RD+E NHS FT. Training will be in accordance with the manufacturer’s User Manual (technique guide) and Instructions for Use (IFU).

The four NHS sites selected to participate in the trial have dedicated knee surgery teams who are experienced in conventional HTO surgery. All sites have the required radiology and professional healthcare support facilities.

##### Specific Procedures for Use

Instructions for Use (IFU) outlines key information relating to the use of the device. The User manual outlines the preparation and the surgical steps required during use (see User Manual).

## JUSTIFICATION FOR THE DESIGN OF THE CLINICAL INVESTIGATION

High tibial osteotomy (HTO) is a well-established and successful surgical procedure. Patient outcomes depend greatly on the accuracy of achieving the correction planned prior to the surgery (27). HTO procedures using personalised devices have not only the potential to reduce irritation compared to generic ‘one size fits all’ plates, but also allow a more accurate surgical correction. The TOKA® product is a novel personalised HTO procedure with the potential to achieve this.

### Pre-clinical Evidence

The TOKA device has been extensively and systematically evaluated to demonstrate safety, effectiveness, and appropriateness for clinical use. The full summary of pre-clinical testing can be found in the TOKA Investigator Brochure.

## RISKS AND BENEFITS OF THE INVESTIGATIONAL DEVICE AND CLINICAL INVESTIGATION

### Anticipated Clinical Benefits

High tibial osteotomy (HTO) is a proven treatment to reduce the pain of knee osteoarthritis. A large body of evidence demonstrates that HTO performed with correct indications and surgical technique has good long-term results. Well-performed HTO can delay the need for knee replacement by 10 years and in some individuals it can be the definitive treatment, thereby avoiding the need for further surgery altogether.

Potential benefits specific to the TOKA® HTO procedure include:

- Personalised implants to reduce the likelihood of soft tissue irritation
- Personalised surgical instrumentation to reduce surgery time and increase surgical accuracy

### Anticipated Adverse Effects

As with all major surgical procedures, side effects and adverse events can occur. While many possible reactions may occur, some of the most common are listed in Table 3.

**Table 3.** Anticipated Adverse Effects.

| Anaesthetic Related | Surgical Procedure Related |
| --- | --- |
| Death | Iatrogenic neural and vascular injury |
| Complications secondary to existing co-morbidity e.g. ischaemic heart disease. | Damage to soft tissues incl. swelling |
| Bleeding intra and post operatively | Abnormal scar formation |
| Pulmonary Embolism | Functional impairment of the musculoskel-etal system |
| Deep Vein Thrombosis | Sudeck's disease |
| Confirmed cerebrovascular accident (CVA) | Allergy/hypersensitivity reactions |
| Confirmed Myocardial Infarction | Side effects associated with hardware prominence* |
| Nausea, vomiting, dental injuries, neurological impairments. | Malunion |
|  | Non-union |
|  | Removal of Tibial Plate |
|  | Fracture of Tibia |
|  | Wound infection |
|  | Septicaemia |
|  | Unexplained knee pain |
|  | Knee stiffness |
|  | Mechanical implant failure* |
|  | Surgical Procedure Related |
|  | Haematoma |
|  | Excessive bleeding |
|  | Above knee amputation |

Complications may result in the need for further surgery such as: Revision Operations, Arthroscopy, Washout, Manipulation under Anaesthetic, Debridement (open) and aspiration.

*These anticipated adverse effects entries are associated with the device.

### Residual Risks associated with the investigational device

Individuals that have a known sensitivity to device materials will be excluded; nevertheless it is possible that a patient could develop a reaction to the device materials. In extreme cases this could require implant removal.

### Mitigation of Risk

All risks have been mitigated as far as possible, an ISO compliant design process was used for developing the ToKa system. Risk control measures are outlined in the Investigator Brochure.

### Risk to Benefit Rationale

A complete Risk Benefit Analysis according to ISO 14971 can be found in the Risk Management File. The final evaluation of the overall residual risk acceptability showed that the medical device was conforming to the requirements set out in the criteria for risk acceptability and that the overall risk was small compared to the beneficial effects that the product could bring (i.e. more accurate correction, decreased invasiveness, return to an active lifestyle, reduction in pain, comparable if not improved adverse events, preservation of natural structures within the knee, and reduced cost).

## OBJECTIVES

The trial objectives are listed in Table 4

**Table 4.**
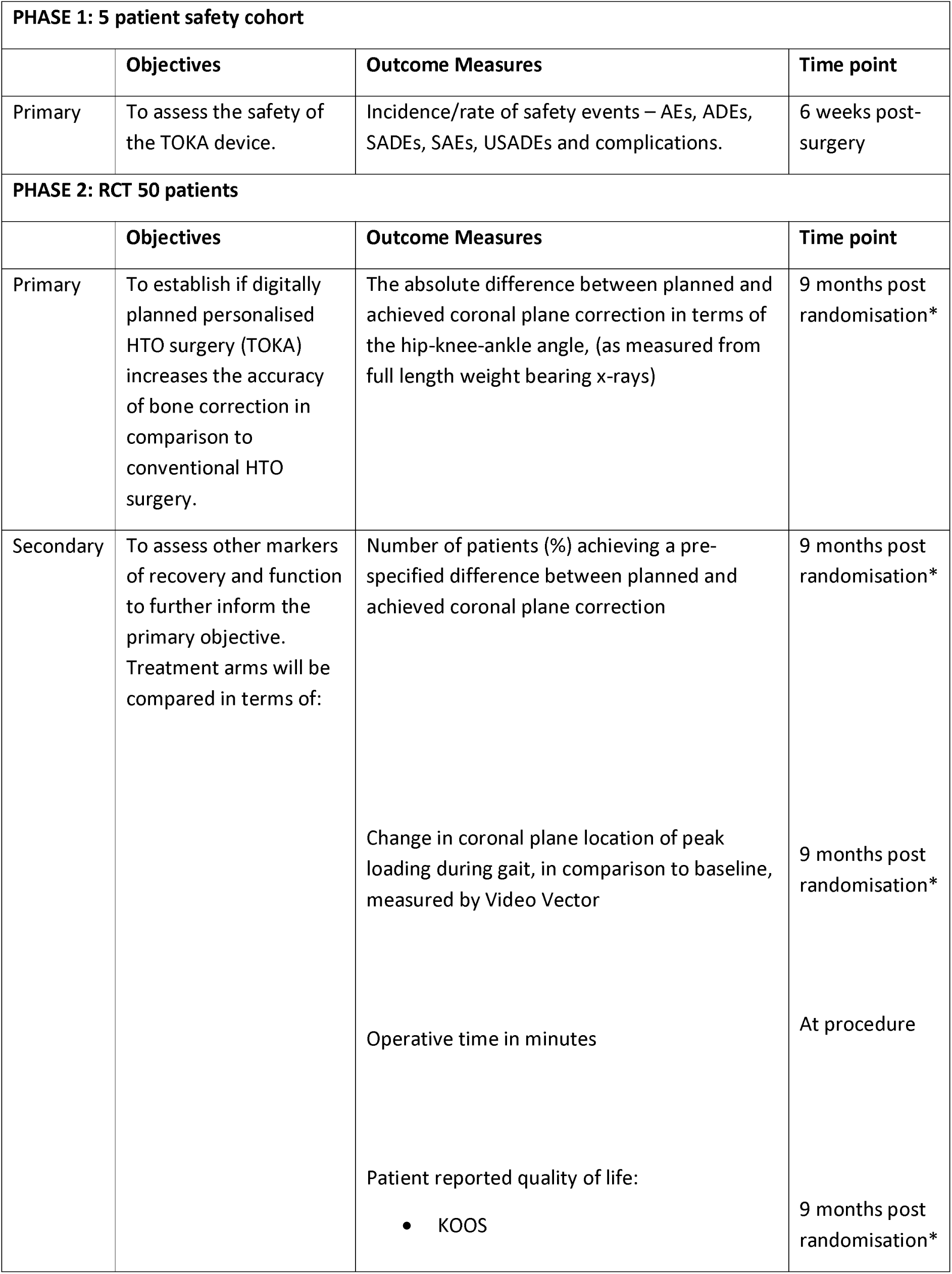
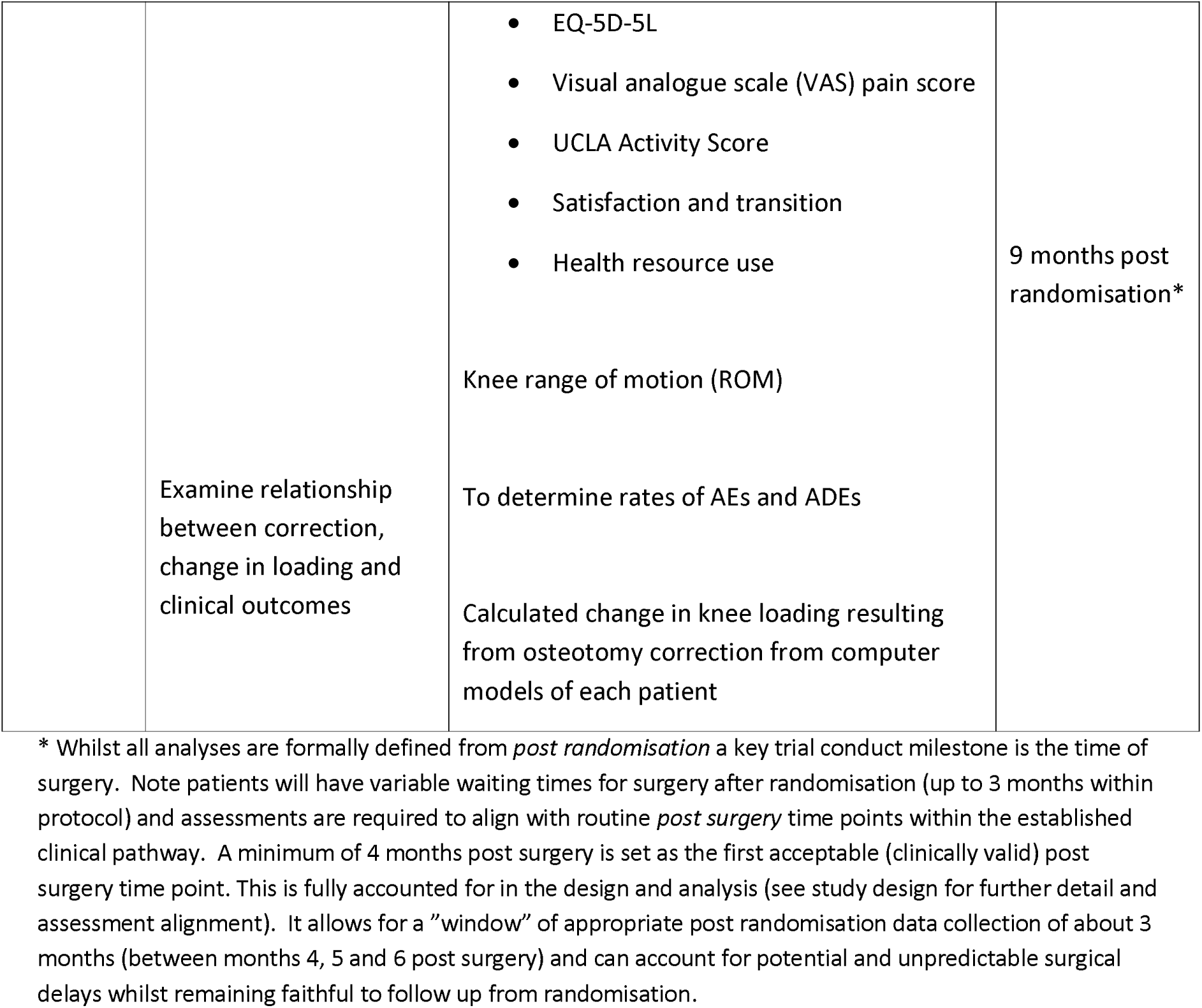
Trial Objectives.

### Primary Outcome Measures

A full leg weight bearing X-Ray will be taken at baseline and at 9 months post randomisation. The images will be used to calculate the absolute difference between planned and achieved coronal plane correction in terms of the hip-knee-ankle angle. Note for all measures 9 months post randomisation are intended to align with the 6 months post-surgery within the standard protocol (surgery within 3 months of randomisation and no delays). These time points can therefore be used interchangeably for the purposes of trial management.

The amount of Hip Knee Ankle (HKA) correction required is decided by the surgeon using landmarks on the weight-bearing X-ray.

The Hip-Knee-Ankle (HKA) angle is defined as the lateral angle between two lines: one line from the centre of the femur head using Mose circles to the middle of the distance between the tibial spines, and a second line from the centre of the ankle to the centre of the tibial spines. An angle of more than 180 degrees denotes a varus alignment.

The baseline HKA angle and the planned HKA correction angle is recorded pre-operatively. The HKA correction angle achieved is recorded post-operatively.

The primary outcome measure compares the planned correction against the actual HKA correction achieved, determined by the difference in degrees, between actual pre-op alignment and actual post-op alignment.

The follow-up X-ray imaging taken 9 months post randomisation will be anonymised and sent to two blinded reviewers to independently measure the difference between planned and achieved correction. If a consensus cannot be reached between the two reviewers, a third blinded reviewer will be consulted.

A service evaluation will be conducted to demonstrate the agreement and repeatability of the measurement of the planned correction, therefore removing the requirement for independent review of the baseline X-ray imaging.

### Secondary Outcome Measures

#### Number of patients achieving a pre-specified difference, 3 degrees, between planned and achieved coronal plane correction

The number of patients who have difference between planned and achieved correction of less than 3 degrees in each treatment arm will be compared at 9 months post randomisation.

#### Change in coronal plane location of peak loading during gait

Video Vector Analysis (VVA) will be used at baseline and at 9 months post randomisation. VVA simultaneously records ground reaction force data from a force plate and video data from a video camera focussed on the lower limb of a participant. This enables the magnitude, orientation, and location of the ground reaction force to be determined relative to the limb under investigation.

#### Operative time in minutes

The difference in operative time (knife to skin – dressings on) and theatre time (into anaesthetic room – out of theatre) between the treatment arms will be compared.

#### KOOS

The Knee injury and Osteoarthritis Outcome Score (KOOS) will be collected at baseline, 9 months post randomisation and 15 months post randomisation. KOOS is a patient reported outcome measure derived from 5 subscales; symptoms (including stiffness), pain, function (daily living), function (sports and recreation activities) and quality of life with scores ranging from 0 – 100, a higher score indicating better health.

#### EQ-5D-5L

To measure quality of life the EQ-5D-5L will be collected at baseline, 9 months post randomisation. The EQ-5D-5L is a validated, generalised, health related quality of life questionnaire consisting of 5 domains related to daily activities with a 5-level answer possibility, which will be converted into multi-attributed utility scores using established algorithms.

#### Visual analogue scale (VAS)

A VAS will be used to measure patient reported pain. Patients will be requested to mark on a scale of 0 (no pain) to 10 (worst possible pain) the number that represents their knee pain at baseline, 9 and 15 months post randomisation. A second VAS score will measure soft tissue irritation at 9 and 15 months post randomisation.

#### UCLA Activity Score

The UCLA Activity score will be collected at baseline, 9 months and 15 months post randomisation. The UCLA Activity Score is a scale ranging from 1 to 10. The patient indicates their most appropriate activity level, with 1 defined as “wholly inactive, dependent on others, and cannot leave residence” and 10 defined as “regularly participates in impact sports”.

#### Satisfaction and Transition

To measure patient satisfaction and transition at 9 months and 15 months post randomisation, participants will be asked how satisfied they are with their knee, how the problems related to their knee compare to before their operation and how willing they would be to have the operation again. General health will also be collected, and the participants will be asked to compare their general health now, to one year ago.

#### Health Resource Use

This study will collect information on participants’ health resource use at 9 months and 15 months post randomisation, including time in operating theatre, visits to primary care, and hospital care services, during the follow-up. Summaries will be presented by trial arm and mean differences with 95% confidence intervals.

#### Range of Motion (ROM)

ROM flexion and extension will be measured (in degrees) at baseline and nine months post randomisation.

#### Change in Knee Loading

Computer models based upon the CT geometry and informed by the achieved correction and video vector analysis will be used to calculate the change in knee loading due to the HTO surgery for each patient. This will enable the relationship between amount of correction, change in loading and outcome to be examined with the aim of optimising HTO surgery.

#### Safety

Safety data will be collected throughout the duration of the trial, to determine the rates of Adverse Events and Adverse Device Effects.

## DESIGN OF THE CLINICAL INVESTIGATION

Pashion is a multi-centre, blinded, superiority two arm, parallel group design randomised controlled trial of personalised versus conventional HTO surgery. Patients will be randomised in a 1:1 ratio. Patients and primary outcome assessors will be blinded to the allocation.

Embedded within the trial is a non-randomised pre-trial technology check and safety assessment, this is termed the Initial Safety Phase (Phase 1).

### Phase 1 – initial safety phase (5 patients)

During this stage of the clinical investigation, 5 patients fulfilling the inclusion criteria will be recruited and assessed in an identical way to the 88 patients recruited in the main trial, but without randomisation. The 5 participants will receive the interventional device – TOKA.

The aim of Phase 1 is to assess the safety of the TOKA device. An independent DSMC will meet 6 weeks after the six-week assessment on the fifth patient is complete, to determine whether the study can proceed to the Randomised Controlled Trial (Phase 2). Although the decision to proceed to Phase 2 is based on the safety of the procedure (an independent review of any intra-operative and short-term post-operative safety data); safety reporting for Phase 1 participants will continue throughout the duration of their study follow up.

### Phase 2 –RCT (50 patients)

Recruitment of the remaining patients (Phase 2) will take place after the six-week assessment on the fifth patient is complete and the oversight committee supports progression to Phase 2. Patients will be randomised to receive personalised (TOKA) or standard HTO.

Patients in Phase 1 and 2 will be followed up in the same manner for a period of up to 15 months from registration (Phase 1) and randomisation (Phase 2) (12 months from surgery). Follow up data will be collected via post or an electronic data collection system whereby patients can complete their patient reported outcome measures (PROMs) directly into a secure on-line portal. Follow up radiological data (X-ray) from patient reports and images will be collected at the standard 6 months post-surgery appointment. Both groups will receive the same standard pre-operative and post-operative routine care.

In addition to routine care, all study participants will undergo a pre-operative CT scan to the affected leg which will be a spiral CT through the hip, knee and ankle, following the established very low dose Imperial CT protocol (28). The participants will also undergo pre-operative video vector analysis, by performing five walking trials over a force platform with synchronised video of the lower limb, allowing the coronal plane location of peak loading to be measured. Full leg weight bearing x-ray, which is the routine imaging technique for HTO planning, will be taken pre-operatively according to the protocol of Brouwer et al (29). To align post-randomisation and post-surgery assessments as much as possible, sites are asked to book follow-up appointments in line with the scenarios outlined in Appendix A.

### Recruiting Centres

Participants will be recruited from four NHS hospitals located across the United Kingdom. The number of recruiting centres may be increased as the clinical investigation progresses.

## EMBEDDED BIOMECHANICAL SUB-STUDY (CARDIFF AND VALE UHB SITE ONLY)

### Description of the Sub-study

Participants who have consented to take part in the PASHiOn RCT at the Cardiff and Vale University Health Board (UHB) site will be invited to participate in a biomechanics sub-study which will involve attending two sessions at the Musculoskeletal Biomechanics Research Facility (MSKBRF) at Cardiff University. The first session will take place approximately 2-4 weeks pre-surgery and the second session 6 months post-surgery.

### Sub-study Aims and Objectives

#### Primary aim

To understand how knee realignment surgery changes the internal movements of the knee and whether this information can be used to improve surgical planning.

#### Secondary Objectives

- To assess the relationship between internal movements of the knee joint as calculated through the primary aim to a number of different methods of assessment of recovery and function including:-

- Patient reported function determined from Oxford Knee Score and Knee Outcome Survey
- Patient reported pain as determined from the Pain Audit Collection System (PACS)
- Changes in coronal plane correction (as determined by hip-knee-ankle angle)
- Muscle activation patterns as measured by Electromyography (EMG).
- To use the movement and contact position information of the knee joint and how this changed post-surgery to improve existing surgical planning tools to eventually improve outcome.

#### Sub-study Design and Methods

This sub-study will run alongside the PASHiOn trial in the Cardiff and Vale UHB site. No additional randomisation of participants will be performed. The randomisation allocations for participants in the main PASHiOn trial will form two blinded groups within the sub-study.

Participants will attend baseline and follow-up visits at the Musculoskeletal Biomechanics Research Facility within Cardiff University School of Engineering. The baseline visit will take place prior to HTO surgery and the follow-up visit will take place around six months post-surgery at the time of the VVA follow-up analysis for the main study. Visits will ideally combine both the main trial VVA and the substudy activities at a single appointment time.

During each visit participants will complete an additional series of Patient Reported Outcome Measures along with those for the main study and their biomechanical movement will be analysed using dynamic x-ray and 3D motion analysis. These data will be combined with x-ray, CT scan and the participant reported outcome data (KOOS, EQ-5D-5L, UCLA Activity Score, Visual Analogue Scale and the 6- and 9-month follow-up questionnaires) that has been collected as part of the PASHiOn trial to allow the exact movement of the knee joint to be mapped.

## Participant Identification

### Study Participants

Alongside obtaining consent for participating in the PASHiOn trial, patients at Cardiff and Vale UHB will also be asked to take part in the sub-study. Participation in the sub-study is optional.

### Inclusion Criteria

To be considered eligible, the participant must meet all of the inclusion criteria as part of the PASHiOn Trial (Section 8.1.1) and the following inclusion criteria:

- Participants are enrolled in the PASHiOn trial in Cardiff.
- Willing and able to give informed consent Exclusion Criteria

### Patients will be considered ineligible if ANY of the following apply

- Self-reported inability to walk 10 meters without the use of a mobility aid.
- If patients have withdrawn from PASHiOn trial
- If patients have already received treatment within the PASHiOn trial
- Unable to comply with the requirements of the study Study procedures

### Recruitment

Any patient who agrees to take part in the PASHiOn trial at the Cardiff and Vale UHB trial site will be provided with a full Patient Information Sheet for the sub-study at the earliest opportunity. This may be sent via post, email or given to the patient at the PASHiOn baseline visit where it has not been possible to provide to the patient at their clinic visit.

Patients will be approached for consent following the completion of their PASHiOn baseline visit. Patients will be given as much time to consider their participation in the study as they require which will be at least 24 hours.

### Screening and Eligibility Assessment

All patients who are deemed eligible to take part in the PASHiOn trial will be screened for inclusion in this study. Screening Logs will be kept to record the number of patients approached to take part in the study. Reasons for decline will be sought, but patients are not obliged to provide these.

Screening will occur approximately four months before the planned date for surgery. Eligibility will be confirmed at the PASHiOn baseline visit around three months before surgery. In order to ensure sufficient time for the baseline data collection in this study consent should be in place no later than one month before the expected surgery date.

### Informed Consent

Procedures for obtaining Informed consent for the sub-study will be followed as described in section 10.2 for the main PASHiOn trial. It will be made clear to patients that agreeing to take part in the PASHiOn trial does not mean that they have to take part in the sub-study. Patients will be given ample time to read the information and ask questions prior to deciding about their participation in the study.

During the consent discussion, patients will be informed of what taking part in the study will involve including the timing and location of pre and post-surgery lab visits and the extra time commitment for the study in addition to the main study. Patients will be made aware of any potential benefits or risks involved with participation in the study.

If the patient is happy to take part in the study they will be asked to complete a written consent form at the end of the PASHiOn baseline visit. If the patient requires more time to consider their participation in the study they will be provided with an option for remote consent.

The unique study ID allocated to each participant for the main trial will be used in the sub-study.

### Randomisation, Blinding and Unblinding

Patients will not be randomised within this study but care will be taken to ensure that patients remain blinded to their PASHiOn trial allocation.

During the post-surgery visit to the fluoroscopy lab all screens which could potentially unblind patients to their treatment allocation will be switched off or turned away from the patient.

If a patient does become unblinded to their PASHiOn allocation during the post-surgery visit this will be reported to the main trial management team in Oxford at the first available opportunity.

### Baseline Assessments

Participant contact information including; full name, date of birth, address, telephone number and email address will be accessed from the PASHiOn study site database. These data will be maintained for the duration of the participant’s involvement in the study to arrange the pre and post-surgery visits to the MSKBRF.

The baseline (Pre-surgery) data collection will take place in the Fluoroscopy laboratory at the MSKBRF. The visit will take around 120 minutes to complete and will be conducted by appropriately trained and delegated members of the MSKBRF Team.

### Baseline (Pre-Surgery) Visit Overview (2-4 weeks pre-surgery)

- Confirmation of consent
- Pre-Assessment screening (as per IR(ME)R requirements)
- Completion of Patient Reported Outcome Measures (PROMs)

- xford Knee Score (OKS)
- Outcome Survey (KOS)
- Audit Collection System (PACS)
- Anthropometric measurements

- Height
- Weight
- Thigh girth
- Waist circumference
- Combined motion capture and bi-plane fluoroscopy assessment (see Section 7.2.8)

### Follow-Up Visits

The post-surgery follow-up visit will take place no earlier than six months after surgery. All follow-up assessments will be completed in the fluoroscopy laboratory at the MSKBRF. Follow-up data collection will take approximately 120 minutes and will be completed by an appropriately trained and delegated member of the MSKBRF team.

6 Month Follow-up Visit Overview (6 months post-surgery)

- Confirmation of continued consent
- Pre-Assessment screening (as per IR(ME)R requirements)
- Completion of Patient Reported Outcome Measures (PROMs) – These may be completed by the patients via post/email in the week before the visit.

- xford Knee Score (OKS)
- Outcome Survey (KOS)
- Audit Collection System (PACS)
- Anthropometric measurements

- Height
- Weight
- Thigh girth
- Waist circumference
- Combined motion capture and bi-plane fluoroscopy assessment (see Section 7.2.8).

## Motion Capture and Fluoroscopy Assessment

Passive retro-reflective markers will be attached to the relevant bony landmarks and the participant will also be asked to stand still for short periods of time (1 second). This allows calibration for the individual participant and establishes both technical and anatomical 3D coordinate axes of the joint. The position information is processed later using specially designed computer software to relate anatomical axes to technical axes using transformation matrices, a neutral, quiet joint position is established.

Small, lightweight, wireless, surface EMG electrodes will be positioned on the lower limbs. These sensors measure muscle activity and work best when placed directly onto the skin, therefore we may request permission to shave and/or exfoliate a small patch of skin. Participants are free to decline this request, and the data collection can still continue if they do so.

The participant will then be asked to perform a number of set knee movements whilst being recorded by both the motion capture cameras and the biplane pulsed x-ray machine. These actions will be directed by either the researcher who is present and who is an IR(ME)R entitled operator or by a trained radiographer at the School of Engineering. This will take approximately 90 minutes where the X-ray exposure will only take up a small amount of that time. The following data collection steps will take place involving the participant although the order of these may change at the appointment:

- Initial static calibration with just motion capture
- Static calibration with combined Biplane X-Ray and Motion Capture (Maybe repeated if required)
- Stair Ascent (up to 5 trials) with combined Biplane X-Ray and Motion Capture
- Stair Descent (up to 5 trials) with combined Biplane X-Ray and Motion Capture
- Static calibration for level gait activity with Biplane X-ray and motion capture (Maybe repeated if required)
- Level gait activity (up to 5 trials) with combined Biplane X-Ray and Motion Capture.
- Static calibration for knee lunge task with combined Biplane X-Ray and Motion Capture (Maybe repeated if required)
- Knee lunge activity (up to 5 trials) with combined Biplane X-Ray and Motion Capture

Some additional trials with just Motion Capture may be undertaken to help familiarise the participant with the task prior to X-Ray and to ensure enough movement trials have been captured. If a participant feels unable to perform a particular movement this will be recorded and data collection will continue.

Participants will be recorded using audiovisual cameras during the sessions. This is to allow re-assessment of results as a quality assurance measure. All data files and audiovisual files will be digitally stored on password protected Cardiff University computer drives. Participants anonymity will be ensured in any video content being presented / published using digital masking methods. Consent will be obtained for the use and storage of video data.

## Additional radiation exposure

Enrolment in the sub-study will result in the participant receiving additional exposure to X-ray radiation. For the knee, the maximum effective radiation dosage has been assessed by the Radiation Protection Service, Velindre NHS Trust, and is estimated to be a maximum of 0.326 mSv (milliSieverts) for two visits that a participant might be invited to attend. This corresponds to just under 2 months of exposure of natural background radiation, to which we are all exposed continuously. Ionising radiation can cause cancer which manifests itself after many years or decades. The risk of developing cancer as a consequence of taking part in this study is 0.0016 %, which is low. For comparison, the natural lifetime cancer incidence in the general population is about 50%. This places this imaging investigation with two visits within the level of risk graded as Category IIa – minor to intermediate, defined in ICRP publication 62 “Radiological Protection in Biomedical Research”.

### PARTICIPANTS

#### Description of Study Participants

Patients aged 18 to 65 years with symptomatic medial compartment osteoarthritis of the knee confirmed using clinical assessment and x-ray.

##### Inclusion Criteria

To be considered eligible, the participant must meet all of the following inclusion criteria:

- Patients undergoing High Tibial Osteotomy
- Male or Female, aged 18 to 65 years
- Primary diagnosis of Non-Inflammatory Degenerative Joint Disease (NIDJD)
- Predominately diagnosed with unicompartmental medial osteoarthritis of the knee with the normal clinically acceptable level of other compartmental involvement
- Varus deformity <20°
- BMI ≤ 35. An exemption to this may be made if the participant (in the investigators opinion) is suitable for surgery.
- Participant is willing and able to give informed consent for participation in the study.
- Able (in the Investigators opinion) and willing to comply with all study requirements.
- Willing to allow his or her General Practitioner and consultant, if appropriate, to be notified of participation in the study.

##### Exclusion Criteria

Patients will be considered ineligible if ANY of the following apply:

- Female participants who are pregnant, lactating or planning pregnancy during the course of the study.
- Prisoners
- Participants with a known deep tissue sensitivity to device materials
- Participants with an active or suspected latent infection in or about the affected knee joint
- Participants who have received any orthopaedic surgical intervention to the lower extremities (excluding investigative surgery) within the past 12 months, or is expected to require any orthopaedic surgical intervention to the lower extremities, other than the HTO to be enrolled in this study, within the next 12 months (including intra-articular procedures).
- Participants who require bilateral HTO with surgery planned on their second knee within 6 months of their first operation (bilateral HTO patients are otherwise included).
- Participants who require bilateral HTO who have a previous unsuccessful contralateral partial replacement or HTO.
- Chronic heart failure (NYHA Stage ≥ 2)
- Neuromuscular or neurosensory deficiency, which limits the ability to evaluate the safety and efficacy of the device
- Systemic disease diagnosis (e.g. Lupus Erythematosus) or a metabolic disorder (e.g. Paget’s disease) leading to progressive bone deterioration.
- Participant is immunologically suppressed or receiving steroids in excess of normal physiological requirements (e.g. > 30 days).
- Participant is a smoker.
- Any other significant disease or disorder which, in the opinion of the Investigator, may either put the participants at risk because of participation in the study, or may influence the result of the study, or the participant’s ability to participate in the study.

## INVESTIGATIONAL DEVICE AND COMPARATOR

The investigational device and comparator are two types of metal plate used to fix the bone in place during a high tibial osteotomy (HTO).

### Justification for use of Comparator

In order to detect a difference in outcome, surgery that includes the use of the personalised device needs to be compared to the current standard treatment. The Tomofix device is available in 2 sizes (standard and small, both with 8 screw holes) and was used in more than 80% of HTO procedures reported upon in the UK Knee Osteotomy Register Annual Report 2018. The Tomofix procedure is similar to the TOKA procedure; however, it is not patient specific and does not use custom-made plates or surgical guides. The ActivMotion system (NewClip Technics, France) is an HTO system with 2 sizes of fixation plate (8 screw holes/large and 6 screw holes/small). The ActivMotion and ActivMotion S devices (NewClip) represented a combined 11% of osteotomy devices used in the UK Knee Osteotomy Register Annual Report 2018 and since then the use of the ActivMotion and ActivMotion S devices has increased significantly. Like the Tomofix device, ActivMotion plates are NOT custom-made however, NewClip offer the option of using custom-made plastic PSI surgical guides. The specific device and plate size used for each participant receiving the comparator will be recorded.

Both groups will receive the same standard pre-operative and post-operative routine care. There are no medications relevant to this clinical investigation. All medication will be prescribed as per routine care.

### Number of Investigational Devices to be Used

The clinical investigation will use up to 49 investigational devices. For each patient receiving the intervention, one device will be used.

#### Total Expected Duration: Clinical Investigation

The clinical investigation is expected to last up to 66 months.

#### Expected Duration: Per-Patient Participation

The time that each patient remains in the clinical investigation will vary depending on surgical waiting times. Participants will be followed up for 6 months (minimum of 4 and maximum of 12 months) from the day of surgery (9 and 15 months from randomisation).

#### Sample Size

A total of 55 patients will be included in the clinical investigation. The sample will be achieved over 2 stages: Phase 1 – initial safety phase (5 patients) and Phase 2 – RCT (50 patients).

The sample size calculation was based only upon the primary outcome of value of absolute difference between achieved and planned correction. A total sample size of 50 (25 per treatment arm) will be sufficient to detect a standardised difference of 0.65, using 80% power, a two-sided 5% significance level and allowing for 10% loss to follow-up. This standardised difference corresponds to a difference of 2.6 degrees, which we deem clinically relevant, and a standard deviation of 3, a conservative estimate based on a recent review of alignment corrections after HTO (23) and a recently published clinical article of a non-randomised trial in Bologna Italy involving 25 recipients of the TOKA plate, in which the overall mean difference between planned and achieved correction in terms of HKA was 2.1° (SD ± 2.0°)(30).

## PROCEDURES

The investigator will assume overall responsibility for study procedures undertaken at site. However, they may delegate tasks to qualified and appropriately trained members of the local PASHiOn team. Delegations will be recorded on the Delegation and Responsibilities Log. It is the responsibility of the local PI to ensure that this log is maintained, reflects the current situation at site and is filed appropriately in the Investigator Site File (ISF).

In response to the Covid-19 pandemic, all participating sites will undertake necessary precautions and adhere to relevant guidelines.

### Participant Screening, Identification and Recruitment

Recruitment will run in two-phases. The first will involve recruiting five patients to the non-randomised Initial Safety Phase (Phase 1). Once safety has been reviewed at 6 weeks and the oversight committee deem it appropriate to do so, recruitment will roll out for the randomised phase of the trial (Phase 2). Randomisation to Phase 2 will be performed once consent has been given and baseline assessments have been completed.

Patients will be identified in outpatient clinics or from surgical waiting lists and will be screened for eligibility by members of the clinical team. Routinely completed X-ray images and reports will be screened by the local care team, to confirm the diagnosis of medial compartment osteoarthritis.

Patients deemed eligible and listed for an HTO procedure will first be approached about the trial by their treating clinician. Local research teams will provide further information about the trial, both written and verbal, ensuring that the patient is aware of which phase of the trial they are taking part in and that they receive the appropriate Patient Information Sheet and Consent Form.

If the clinician is able to confirm eligibility prior to an appointment, an invitation letter and a Patient Information Sheet (PIS) will be sent to the patient ahead of their appointment. The clinician will re-confirm eligibility and willingness to participate with the patient at the appointment and then refer them to the local clinical research team (for example, the local research nurse) for recruitment and arrangement of screening and baseline assessments.

General information about the number of patients approached who are eligible but decline to participate will be recorded on screening logs at each site. Reasons for decline will be sought, but patients are not obliged to provide these. Information on any patient who meets the inclusion criteria but is then deemed ineligible (meet s an exclusion criteria) will also be recorded on the screening log.

### Informed Consent

Phase 1 and Phase 2 will have separate versions of the Participant Information Sheet (PIS). The Informed Consent Form (ISF) will request confirmation of which phase of the trial the participant is being recruited to. Written and verbal versions of the PIS and ISF will be presented to the participants detailing no less than; the exact nature of the clinical investigation; what it will involve for the participant; the implications and constraints of the Clinical Investigation Plan; the known side effects and any risks involved in taking part. It will be clearly stated that the participant is free to withdraw at any time for any reason without prejudice to future care, without affecting their legal rights and with no obligation to give the reason for withdrawal.

The participant will be allowed as much time as wished to consider the information, and the opportunity to question the Investigator, their GP or other independent parties to decide whether they will participate in the clinical investigation. Written Informed Consent will then be obtained by means of participant dated signature and dated signature of the person who presented and received the Informed Consent. The person who receives consent must be suitably qualified and experienced, and have been authorised to do so by the Principal Investigator. A copy of the signed ISF will be given to the participant. The original signed form will be retained at the investigating centre and filed in the ISF.

Once Consent has been taken, the patient should be immediately listed for a CT scan, which is needed to manufacture the TOKA device.

### Baseline Assessment

The Baseline assessment will be undertaken once the patients’ eligibility and willingness to participate has been confirmed and the patient has provided Informed Consent. No study specific activities will take place prior to receiving Informed Consent.

Baseline assessment includes:

- X-ray (routine imaging to confirm initial diagnosis can be used). In some cases, as per routine practice, this may require a second X-ray to review clinical status of the knee joint if the period from initial X-Ray is more than 6 months.
- CT scan (approximately 3 months prior to surgery). Dependent on unexpected delays to surgery, a repeat CT scan may need to be undertaken. The need for a repeat scan will be a clinical decision, but with a maximum window of 6 months between the scan and surgery.
- Video Vector Analysis
- Range of Motion (standard clinical measurement)
- Pregnancy test (standard of care)

The baseline assessment includes a questionnaire to be completed by the patient which will take approximately 20 minutes. These include:

- KOOS
- EQ-5D-5L
- Pain VAS
- UCLA Activity Score

General patient demographics and patient contact details will also be collected to help co-ordinate the follow up assessments.

Please refer to Appendix C for a detailed flow of the trial investigations and approximate timings.

## Randomisation, Blinding and Unblinding (relevant to Phase 2 only)

### Randomisation

Randomisation to Phase 2 will be undertaken using a centralised web-based randomisation service (RRAMP) run through the Oxford Clinical Trials Research Unit (OCTRU). Randomisation will take place once the CT imaging is available, approximately 3 months prior to the HTO operation when the planned date of surgery is confirmed. This will allow sufficient time for the plates and surgical guides to be manufactured, delivered to the appropriate study site and sterilised. Once a patient has been randomised, the CT scan, X-ray imaging and planned correction angle for participants allocated the intervention arm (TOKA), will be securely sent to the device manufacturer. A data transfer guidance document detailing the transfer procedure will be supplied to all recruiting sites. The local site clinician will need to be available to assist with the design of the device, to ensure that there are no delays with manufacture.

Consenting participants will be randomised at clinic on a 1:1 basis to either personalised or standard HTO. Random allocation will be implemented using a minimisation algorithm stratified by trial centre participant sex, age and BMI. The minimisation algorithm will include a random element to prevent predictability of the treatment allocation. A small number of participants will be randomised using a simple randomisation schedule, generated in advance by the trial statistician, the seed the minimisation algorithm. Stratification by centre, sex, age and BMI will help ensure that any related effects will be equally distributed in the trial arms. We have chosen to separate patients younger than 50 years and 50 years and older, and patients with a BMI lower than 30 and those with a BMI of 30 or greater, as stratification cut-off points for this trial.

### Blinding

All patients and outcome assessors will be blinded to the treatment allocation. All patients will have the same pre-operative CT scan to ensure that they are blinded to allocation arm. The radiological assessment will mask the device on the x-ray image, prior to independent measurement of the correction, ensuring that the primary outcome measurement is blinded to the study arm allocation.

### Unblinding

If patient unblinding is deemed necessary, it will be discussed on a case-by-case basis with the central study team and local PI. All unblinding will be at the discretion of the local investigators, when clinically indicated for the safety of the patient.

### Pre-Surgical Assessment

There are no study specific pre-surgical assessments. On the day of surgery, an appropriately qualified member of the local PASHiOn team should meet with the patient to answer any questions, address any concerns and confirm that patient consent is still valid. A record of this contact should appear in the patient notes.

### Intervention

All patients will receive a high tibial osteotomy (HTO) as a single surgical intervention. A high tibial osteotomy is a surgical procedure to realign the leg. A cut is made in the tibia and the alignment of the tibia is carefully adjusted by levering open this cut in the bone until the desired alignment is reached. The bone is then fixed with a plate that is held in place with screws. This is called an ‘opening wedge osteotomy’. The osteotomy gap in the bone will fill in with new bone over the next few months post-surgery. Intraoperative X-rays may be taken as part of routine care. The need for an Intraoperative X-ray will be a clinical decision, and the number of X-rays taken will be recorded on the operative CRF.

### Follow up Assessments

Following surgery, participants will return to hospital as part of routine clinical care at 6 weeks, 3 months and 6 months post-surgery. No data will be collected for the study at the 6 week and 3 month visits but recruiting centres will be expected to routinely monitor patient records at each post-operative time point, for unreported safety events that may have occurred.

A full leg weight bearing X-Ray will be taken as part of routine care at the 6 month post-surgery follow up appointment (9 months from randomisation). Images and reports from this routine scan will be securely sent to the central study team at Oxford, who will ensure that the images are anonymised and masked, before sending to the blinded independent assessors to measure and assess the primary outcome. A repeat Video Vector Analysis will be arranged for 6 months post-surgery (9 months post-randomisation).

The follow up questionnaires will be sent 6 and 12 months post-surgery (approximately 9 and 15 months post randomisation). Participants will have a choice whether they would like to complete these electronically (via a link sent in an email) or completed on paper form sent out via post. The questionnaires will take between 20 and 30 minutes to complete. If the patient does not complete the questionnaire within 14 days of the due date, another questionnaire will be sent. Following this, if we have still not received a completed questionnaire within a further 14 days, a phone call to the patient may be made to complete the questionnaire. If a participant chooses to receive the questionnaires electronically, but then does not complete them, the questionnaires may be posted to them as an additional attempt to collect the data.

The current COVID 19 situation may have an effect on longer term secondary outcome collection (duration of the grant). It is anticipated all outcomes at the primary endpoint can be collected within the current study duration and funding. If needed an extension will be sought but failure to secure this will not compromise the main research questions of the study.

### Discontinuation/Withdrawal of Participants

In consenting to participate in the trial, participants are consenting to the intervention, assessments, follow up and data collection. Recruitment to Phase 1 and randomisation to Phase 2 will only be undertaken once the patient has been informed of all the trial obligations and of the importance of data collection and when the patient has given consent. Each participant has the right to withdraw from any aspect of the trial at any time. In addition to participant self-withdrawal, an investigator may decide to withdraw a participant if considered necessary for any reason including ineligibility either arising during the study or retrospectively, having been overlooked at screening. A withdrawal form will need to be completed.

### Withdrawal from Intervention

If patients have been recruited into the study and then do not receive the intervention or opt to receive the standard implant and not personalised, either due to self-withdrawal or on the recommendation of their clinician, the reasons for withdrawal or change in treatment will be recorded if available, and sites should explain the importance of patients remaining on the trial follow up. If participants are willing, they will be followed up accordingly. PROMs data would be collected; however, the routine X-ray taken 9 months post-randomisation (approximately 6 months post-surgery) would not be undertaken if the patient has not undergone surgery.

### Withdrawal from Follow-up

Participants may withdraw from the follow-up regime or from the trial altogether. If so, their decision should be recorded in the CRF and medical notes and only data up to the point of withdrawal will be collated and analysed accordingly. The patients will be encouraged to discuss treatment options with their clinician.

The local investigator should attempt to contact any patient requesting withdrawal and appropriate care should continue outside of the clinical investigation. A decision to decline consent or withdraw will not affect the standard of care the patient receives.

### Source Data

Source documents are original documents, data, and records from which participants’ CRF data are obtained. These include, but are not limited to, hospital records (from which medical history and previous and concurrent medication may be summarised into the CRF), clinical and office charts, laboratory and pharmacy records, diaries, microfiches, radiographs, and correspondence.

CRF entries will be considered source data if the CRF is the site of the original recording e.g. there is no other written or electronic record of data). A list of applicable CRFs will be identified in the Data Management Plan held centrally.

All documents will be stored safely in confidential conditions. On all clinical investigation-specific documents, other that the signed consent form, the participant will be referred to by the clinical investigation participant number/code, not by name.

### Definition of the End of Clinical Investigation

The end of the clinical investigation is the date of the collection of the last outcome measure.

## STATISTICAL CONSIDERATIONS

### Description of Statistical Methods

A separate statistical analysis plan (SAP) with full details of all statistical analysis planned for the data in this study will drafted early in the trial and finalised prior to any primary outcome analysis. The SAP will be reviewed and receive input from the Trial Steering Committee (TSC) and Data and Safety Monitoring Committee (DSMC). Any changes or deviations from the original SAP will be described and justified in the protocol, final report and/or publications, as appropriate. It is anticipated that all statistical analysis will be undertaken using Stata (StataCorp LP, www.stata.com) or another well-validated statistical package.

A single formal analysis is planned for this trial, which will take place after the final follow-up assessment has been completed and sufficient time allowed for data collection and data cleaning. Analyses will be based on the intention-to-treat principle (participants analysed in allocated groups regardless of actual treatment received), though participants who did not undergo any HTO may be excluded, but will be repeated for the per-protocol population as a sensitivity analysis to test the robustness of the results.

Standard descriptive statistics will be used to describe the demographics between the treatment groups reporting means and standard deviations or medians and interquartile ranges as appropriate for continuous variables, and numbers and percentages for binary and categorical variables. All comparative outcomes will be presented as summary statistics and reported together with 95% confidence intervals; with all tests carried out at a 5% two-sided significance level.

The primary endpoint, i.e. the absolute difference between achieved and planned correction at nine months post-randomisation (six months post-surgery), will be analysed using a linear regression model adjusted for the randomisation variables (centre, sex, age and BMI), baseline misalignment and other baseline prognostic factors, if appropriate. Results from this model will be presented as adjusted mean difference in the absolute difference between achieved and planned correction (termed as “misalignment” from target value) between the groups and corresponding confidence intervals.

Given the overall size of the study no formal subgroup analyses are planned. Missing data pattern will be presented, and sensitivity analyses for missing data will be performed, including missing not random assumptions.

The key secondary outcome, i.e. number of patients (%) achieving a pre-specified difference between planned and achieved coronal plane correction at nine months post-randomisation (six months post-surgery), will be will be analysed by logistic regression model adjusted in line with the primary analysis. The results of this analysis will be presented as adjusted odds ratio and corresponding 95% confidence intervals. The unadjusted risk differences will also be provided to ensure that both relative and absolute effect sizes are presented. All other secondary outcomes will be analysed using generalised linear models adjusted for the stratification factors and other important baseline prognostic factors.

### Criteria for the Termination of the Clinical Investigation

After Phase 1, the independent DSMC will review the safety data for the 5 patients recruited. Based on the safety data presented, the DSMC will make a decision as to whether the trial can proceed to Phase 2.

During Phase 2, the DSMC will review the safety and efficacy data and provide recommendations to the TSC. Both the DSMC and TSC will evaluate the risk of the trial continuing and take appropriate action where necessary.

### Procedure for Accounting for Missing, Unused, and Spurious Data

Throughout the clinical investigation, missing data occurrence will be minimised through careful study design and data management, training of recruiters and site staff, as well as interaction with participants.

Missing data will be described with reasons given where available, the number and percentage of individuals in the missing category will be presented by treatment arm. All data collected on data collection forms will be used, since only essential data items will be collected. No data will be considered spurious since all data will be checked and cleaned before analysis.

The nature and mechanism for missing variables and outcomes will be investigated. Sensitivity analyses will be undertaken assessing the underlying missing data assumptions. Any imputation techniques will be fully described in the SAP.

### Procedures for Reporting and Deviation(s) from the Original Statistical Analysis Plan

A detailed statistical analysis plan will be drawn up prior to patient recruitment or early in the clinical investigation with review and appropriate sign-off following OCTRU SOPs. Any changes to the statistical analysis plan during the clinical investigation will be subject to the same review and sign-off procedure with details of changes being included in the new version. Any changes/deviations from the SAP will be described and justified on the clinical investigation plan and/or in the final report, as appropriate.

## DATA MANAGEMENT AND SHARING PLAN

A study specific participant number and/or code will be used to identify the participants. The name and any other identifying details will not be included in any study data electronic file. Baseline X-ray and CT scans will contain identifiable information such as the name, date of birth, NHS number and study number of the participant as this needs to be linked up with NHS patient records to link with the personalised implant at the time of surgery. Once transferred to the manufacturer, the images will have all personal identifiable information redacted. Follow up X-ray scans will be anonymised, containing only the participants study number. All other patient related data transferred between the main study team and participating centres will be identifiable only with the patient’s unique study number. When identifiable information is transferred to the study team, secure measures such as Sharepoint electronic file sharing, registered post, courier, or secure-mail accounts will be utilised. For quality control reasons, the main study team may initiate monitoring of site files and data collection forms.

All data will be processed according to the current UK Data Protection Act 2018 and UKGDPR and all documents will be stored safely in confidential conditions. A Data Management and Sharing Plan will be produced for the clinical investigation and will include reference to confidentiality, access and security arrangements. All clinical investigation specific documents, except for the signed consent form and follow-up contact details, will refer to the participant with a unique study participant number/code and not by name. Participants’ identifiable data will be stored in accordance with OCTRU SOPs. All trial data will be stored securely in locked cabinets in offices only accessible by swipe card by the central coordinating team staff in Oxford and authorised personnel.

### Access to Data

Direct access will be granted to authorised representatives from the sponsor, host institution and the regulatory authorities to permit clinical investigation-related monitoring, audits and inspections.

## DEVICE ACCOUNTABILITY

Access to the investigation device, TOKA, will be controlled and the device will only be used for the purpose of the clinical investigation and in accordance with this clinical investigation plan.

The central study team will keep records to document the physical location of all TOKA devices, from shipment to the investigation site until use or destruction.

The Principal Investigator or an authorized designee shall keep records documenting the receipt, use or destruction of the TOKA device, including the date of receipt, unique device code, date of surgery and patient identification number.

### Maintenance and Storage of Device

Dependent on the standard practice and/or preference at each centre, the TOKA device will be stored at room temperature. Temperature Logs will not be required.

Once opened, the device must be used in sterile operating theatres.

The TOKA device packaging will be clearly marked as ‘Exclusively for Clinical Investigation’. The PI will be responsible for controlling access, ensuring the device is stored securely and any use is within the constraints of the PASHiOn clinical investigation.

### Compliance with Investigative Device

The investigational device is a single-use implant. As such, participant compliance with the device is not relevant. If, for any reason, the randomised treatment (device) is not used this will be recorded.

## QUALITY CONTROL AND QUALITY ASSURANCE PROCEDURES

The clinical investigation will be conducted in accordance with the current approved clinical investigation plan, GCP, relevant regulations and standard operating procedures.

A monitoring plan will be drafted in line with the risk assessment for the trial and in accordance with GCP standards stated in ISO 14155. Data will be evaluated for compliance with the clinical investigation plan and accuracy of source documents. For any monitoring activity, the Monitor will verify that the clinical investigation is conducted and data are generated, documented and reported in compliance with the clinical investigation plan, GCP and other applicable requirements.

Monitoring activities will be performed according to the current standard operating procedure and clinical investigation monitoring plan.

The clinical investigation will be overseen by independent steering and data/safety monitoring committees.

## SAFETY REPORTING

### Definitions

The definitions of safety reporting criteria are given in table 5

**Table 5.** Safety Reporting Definitions.

| Term | Definition |
| --- | --- |
| **Medical Device (MD)** | Any instrument, apparatus, appliance, software, material or other article, whether used alone or in combination, together with any accessories, including the software intended by its manufacturer to be used specifically for diagnostic and/or therapeutic purposes and necessary for its proper application, intended by the manufacturer to be used for human beings for the purpose of:<br>• diagnosis, prevention, monitoring, treatment or alleviation of disease,<br>• diagnosis, monitoring, treatment, alleviation of or compensation for an injury or handicap,<br>• investigation, replacement or modification of the anatomy |
|  | or of a physiological process,<br>• control of conception,<br>and which does not achieve its principal intended action in or on the human body by pharmacological, immunological or metabolic means, but which may be assisted in its function by such means. |
| **Investigational Medical Device (IMD)** | Medical device being assessed for safety or performance in a clinical investigation.<br>*Note: This includes medical devices already on the market that are being evaluated for new intended uses, new populations, new materials or design changes.* |
| **Adverse Event (AE)** | Any untoward medical occurrence, unintended disease or injury or any untoward clinical signs (including an abnormal laboratory finding) in participants, users or other persons, whether or not related to the investigational medical device.<br>*Note 1: This includes events related to the investigational medical device or the comparator and events related to the procedures involved (any procedure in this Clinical Investigation Plan).*<br>*Note 3: For users or other persons, this is restricted to events related to the investigational medical device and not the procedure involved.* |
| **Adverse Device Effects (ADE)** | Adverse event related to the use of an investigational medical device.<br>*Note: This includes any adverse event resulting from insufficiencies or inadequacies in the instructions for use, the deployment, the implantation, the installation, the operation, or any malfunction of the investigational medical device.*<br>*This includes any event that is a result of a use error or intentional misuse of the investigational medical device.* |
| **Device Deficiency (DD)** | Inadequacy of a medical device related to its identity, quality, durability, reliability, safety or performance, such as malfunction, misuse or use error and inadequate labelling. |
| **Serious Adverse Event (SAE)** | Adverse event that:<br>○ Led to death,<br>○ Led to serious deterioration in health that either: |
|  | • resulted in a life-threatening illness or injury, or<br>• resulted in a permanent impairment of a body structure or body function, or<br>• required in-patient hospitalisation or prolonged the existing hospitalisation, or<br>• resulted in the medical or surgical intervention to prevent life-threatening illness or injury or permanent impairment to a body structure or a body function<br>○ Led to foetal distress, foetal death or a congenital abnormality or birth defect.<br>*Note: A planned hospitalisation for pre-existing condition, or a procedure required by the Clinical Investigation Plan, without a serious deterioration in health, is not considered to be a SAE.* |
| **Reportable SAEs** | The following SAEs are classified as 'Reportable SAEs':<br>○ Any SAE.<br>○ Any Investigational Medical Device Deficiency that might have led to a SAE if:<br>• suitable action had not been taken, or<br>• intervention had not been made, or<br>• circumstances had been less fortunate.<br>○ New findings/updates in relation to already reported events. |
| **Serious Adverse Device Effect (SADE)** | Adverse device effect that has resulted in any of the consequences characteristic of a serious adverse event. |
| **Unanticipated Serious Adverse Device Effect (USADE)** | Serious adverse device effect which by its nature, incidence, severity or outcome has not been identified in the current version of the PASHiOn Investigator Brochure.<br>NOTE: Anticipated: an effect which by its nature, incidence, severity or outcome has been identified in the PASHiOn Investigator Brochure |

### Recording and Reporting Adverse Events (AEs) and Adverse Device Events (ADEs)

During Phase 1 (and for 6 weeks after the fifth patient has had their surgery) all adverse events (AEs), in participants, either observed by the investigator or reported by the participant and whether or not attributed to the device under investigation will be recorded using the current trial specific AE Report Form. Requirements for AE reporting beyond 6 weeks post-surgery for phase 1 patients, and for phase 2 participants, will be agreed by the DSMC and the CIP will be updated accordingly. In addition, all AEs related to the use of the device by the users will also be recorded and the site asked to report to the CTU.

The PI at site, or designee, is responsible for the identification of any AE as defined in the protocol. Each AE must be assessed for seriousness, severity, causality and expectedness.

It will be left to the Investigators clinical judgement to decide whether or not an AE is of sufficient severity to require the participant’s removal from treatment. A participant may also voluntarily withdraw from treatment due to what he or she perceives as an intolerable AE. If either of these occur, the participant must undergo and end of investigation assessment and be given appropriate care under medical supervision until symptoms cease, or the condition becomes stable.

Where an AE results in participant withdrawal during the clinical investigation, or the AE is still present at the end of the clinical investigation, follow up should continue until a satisfactory resolution occurs.

Participants should be encouraged to report any new adverse event that they, or their GP, feels might reasonably be related to the investigational device. This information should be recorded in the source documents, considered and, where necessary, reported as appropriate.

Recruiting centres will be expected to routinely monitor patient records for any events that may have been missed.

### Recording and reporting of Serious Adverse Events (SAEs)

All SAEs must be reported by the PI or designee to the CTU immediately, but no later than 72 hours after the site learn of its occurrence using the SAE report Form within the study database.

The central coordinating team will perform an initial check on SAE reports, request any additional information, and ensure it is reviewed in a timely manner and according to the relevant SOP on safety reporting for investigational medical devices.

### Events will be followed up until resolution

All SAEs will be reported to all Principal investigators (PIs) and all relevant safety information will be reported to the DSMC, according to written procedures.

### Recording and reporting of Device Deficiencies (DDs)

All DDs must be reported by the PI or designee to the CTU immediately, but no later than 72 hours after the site learn of its occurrence using the Device Deficiency Report Form within the study database.

### Assessment of Seriousness

The PI should make an assessment of seriousness as against the standard definition as outlined in section 15.1.

### Assessment of Severity

An assessment of severity is required (mild, moderate, severe, life threatening, death) to determine if an event is at a severity not usually seen. The investigator, or designee, should make an assessment of severity for each event according to their clinical judgement and knowledge of the participant. This will be recorded in the AE and SAE electronic case report form (eCRF). This information is given in table 6.

**Table 6.**
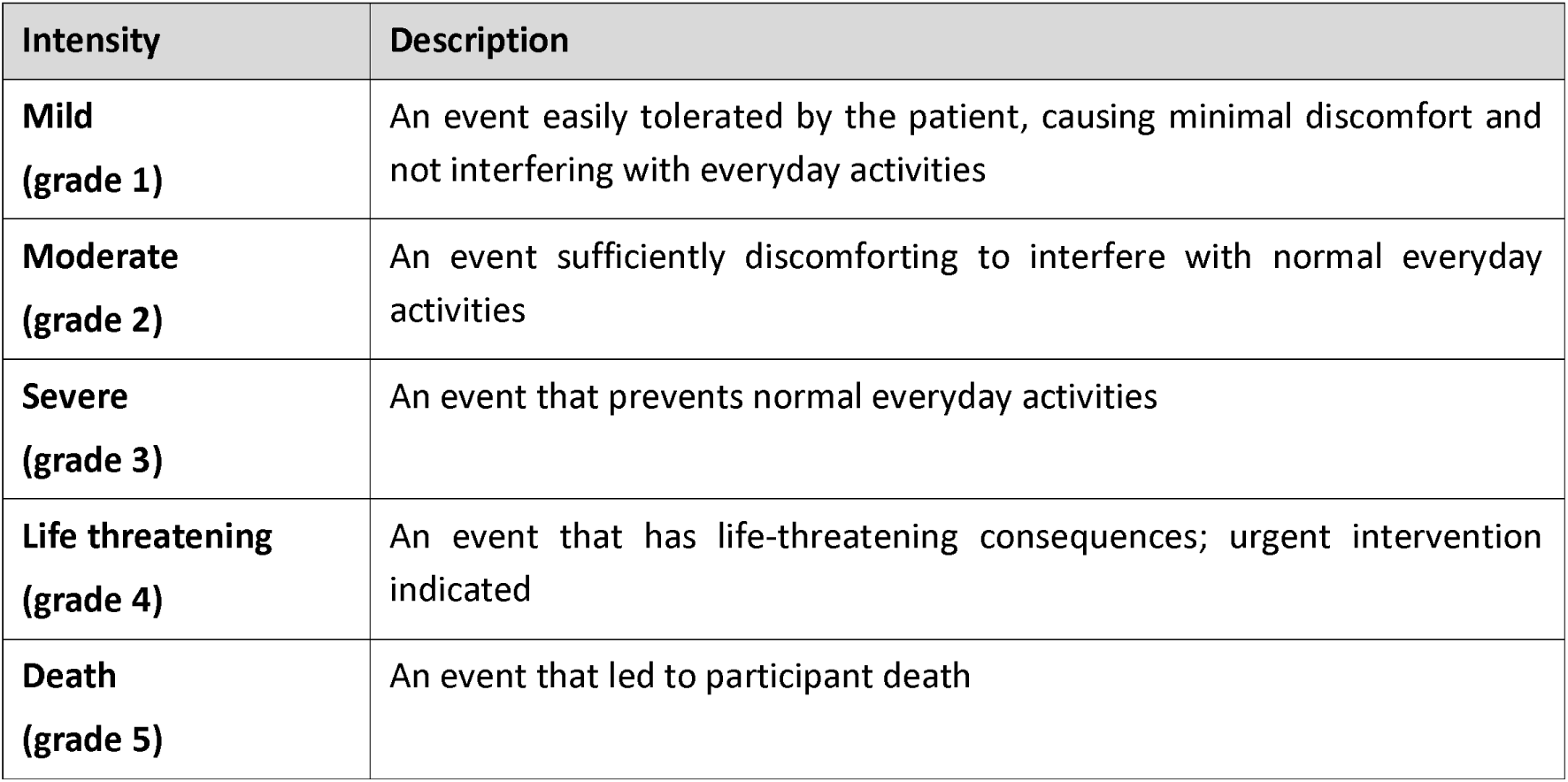
Assessment of Severity.

### Assessment of Causality

The relationship between the use of the investigational medical device (including the medical-surgical procedure) and the occurrence of the adverse event must be assessed by the PI or designee using clinical judgement to determine the causal relationship. Each AE should be categorised as shown in table 7:

**Table 7.** Assessment of Causality.

| Categorisation | Categorisation Description |
| --- | --- |
| **Not Related**<br>Relationship to the device or procedures can be excluded when: | ○ the event is not a known side effect of the product category the device belongs to or of similar devices and procedures;<br>○ the event has no temporal relationship with the use of the investigational device or the procedures;<br>○ the serious event does not follow a known response pattern to the medical device (if the response pattern is previously known) and is biologically implausible;<br>○ the discontinuation of medical device application or the reduction of the level of activation/exposure - when clinically feasible - and reintroduction of its use (or increase of the level of activation/exposure), do not impact on the serious event;<br>○ the event involves a body-site or an organ not expected to be affected by the device or procedure;<br>○ the serious event can be attributed to another cause (e.g. an underlying or concurrent illness/ clinical condition, an effect of another device, drug, treatment or other risk factors);<br>○ the event does not depend on a false result given by the investigational device used for diagnosis, when applicable;<br>○ harms to the subject are not clearly due to use error;<br>**Note 1:** In order to establish the non-relatedness, not all the criteria listed above might be met at the same time, depending on the type of device/procedures and the serious event.<br>**Note 2:** Complications of procedures are considered not related if the said procedures would have been applied to the patients also in the absence of investigational device use/application. |
| **Unlikely** | The relationship with the use of the device seems not relevant and/or the event can be reasonably explained by another cause, but additional information may be obtained. |
| **Possible** | The relationship with the use of the investigational device is weak but cannot be ruled out completely.<br>Alternative causes are also possible (e.g. an underlying or concurrent illness/ clinical condition or/and an effect of another device, drug or treatment).<br>Cases where relatedness cannot be assessed or no information has been obtained should also be classified as possible. |
| **Probable** | The relationship with the use of the investigational device seems relevant and/or the event cannot reasonably explained by another cause, but additional information may be obtained. |
| **Causal Relationship** | The serious event is associated with the investigational device or with procedures beyond reasonable doubt when:<br>○ the event is a known side effect of the product category the device belongs to or of similar devices and procedures;<br>○ the event has a temporal relationship with investigational device use/application or procedures;<br>○ the event involves a body-site or organ that<br>• the investigational device or procedures are applied to;<br>• the investigational device or procedures have an effect on;<br>○ the serious event follows a known response pattern to the medical device (if the response pattern is previously known);<br>○ the discontinuation of medical device application (or reduction of the level of activation/exposure) and reintroduction of its use (or increase of the level of activation/exposure), impact on the serious event (when clinically feasible);<br>○ other possible causes (e.g. an underlying or concurrent illness/ clinical condition or/and an effect of another device, drug or treatment) have been adequately ruled out;<br>○ harm to the subject is due to error in use;<br>○ the event depends on a false result given by the investigational device used for diagnosis, when applicable;<br>**Note:** In order to establish the relatedness, not all the criteria listed above might be met at the same time, depending on the type of device/procedures and the serious event. |

During the causality assessment, clinical judgement should be used and any relevant documents such as current versions of the CIP, IB or risk analysis report should be consulted. The presence of confounding factors such as concomitant medication/treatment, the natural history of the underlying disease, other concurrent illness or risk factors should also be considered.

Investigators will make every effort to collect enough data to adequately define and categorise the event. Situations where the event cannot be properly assessed due to insufficient or contradictory data should be avoided. Where uncertainty remains over the classification of an event, relatedness should not be excluded and the event will be classed as ‘possible’.

The above considerations also apply to adverse events occurring in the comparison group.

Assessment will distinguish between adverse events related to the investigational device and those related to the procedures (any procedure specific to the clinical investigation). An adverse event can be related both to procedures and the investigational device. Complications of procedures are considered not related if the procedure would have been used during standard care i.e. in the absence of the investigational device.

### Assessment of Expectedness

All AEs deemed ‘serious’ and ‘device and procedure related’ (SADEs), must be assessed to determine if the event is ‘anticipated’ or ‘unanticipated’ by the PI or designee. Anticipated Adverse Device Effects are listed in this clinical investigation plan and the Investigator Brochure.

### Expedited Reporting by Sponsor

Expedited reporting of SADEs, USADEs and DDs to the MHRA and Research Ethics Committee (REC) is required by the Sponsor in accordance with the information given in table 8:

**Table 8.** Expedited Reporting by Sponsor.

| Regulatory Body | Event Description | Recording and Reporting |
| --- | --- | --- |
| **MHRA** | The following events are considered as requiring expedited reporting to the MHRA by the Sponsor:<br>• All SAEs that are deemed to be either device related or possibly device related (SADEs/USADEs)<br>• Any device deficiency (DD) that might have led to an SAE if:<br>a) Suitable action had not been taken<br>b) Intervention had not been made<br>c) If circumstances had been less fortunate<br>• New findings/updates in relation to already expedited reported events | For all reportable events where there is an imminent risk of death, serious injury, or serious illness and that requires prompt remedial action for other patients/subjects, users or other persons or a new finding in relation to such events, the Sponsor will report to the MHRA immediately, but no later than **2 calendar days** after Sponsor become aware of such an event or new information in relation to an already reported event.<br>Any other reportable events as outlined above or any new finding/update in relation to them must also be reported immediately, but no later than **7 calendar days** after the Sponsor becomes aware of them. |
| **REC/HRA** | Any SAE/SADE **related** to the administration of the medical device or study procedures and **unexpected** (i.e. not listed in Risk Assessment or Protocol as an expected occurrence) | Will be reported to the REC/HRA by the Sponsor within **15 calendar days** of Sponsor becoming aware of the event using the 'non-CTIMP safety report to REC form'. Any relevant follow-up information should be sought from site and reported by Sponsor as soon as possible after the initial report. |
|  | All SAEs/SADEs/DDs that are reported to Sponsor | A report on the safety of participants will be included in the **annual** progress report submitted by SITU, including Investigator notifications of any SAEs/SADEs/DDs that are reported. Document evidence of investigator review of these will be filed in the Investigator Site File. |

### Deviation from the CIP

The investigator is not permitted to deviate from the clinical investigation plan, except where it is necessary to eliminate an immediate hazard to clinical investigation participants. Under emergency circumstances, deviations from the CIP to protect the rights, safety and wellbeing of participants may be implemented without prior approval from the sponsor, REC and MHRA.

The site must report all deviations to the CTU within 3 days.

The CTU will notify the sponsor, REC and MHRA of any event where it is necessary to deviate from the clinical investigation plan, the nature of and reasons for the deviation. This information will also be recorded in the TMF.

### Serious Breaches

It is the responsibility of the CI to ensure that the trial is run in accordance with GCP and the protocol. This task may be delegated to a suitably qualified or experienced member of the research team but the CI and PI will retain overall responsibility.

A serious breach is defined as “A breach of GCP or the trial protocol which is likely to affect to a significant degree – the safety or physical or mental integrity of the subjects of the trial; or the scientific value of the trial”.

In the event that a serious breach is suspected, the Sponsor must be contacted within 24 hours of identification. Where a serious breach has been identified, it is the responsibility of the Sponsor to notify the REC and MHRA within 7 calendar days of determining that a serious breach has occurred.

### Notification of Pregnancy

Pregnancy will be tested as standard of care to confirm the eligibility of patients for surgery and therefore recruitment to the trial. During follow up, if a participant becomes pregnant, the related site must complete a Pregnancy Notification Form and submit this to the central study office in Oxford. If the pregnancy is prior to the routine X-ray undertaken 9 months post-randomisation, the X-ray will not be performed as per standard of care. All other data (including PROMs) would still be collected.

## URGENT SAFETY MEASURE (USM)

An Urgent Safety Measure (USM) is an action that the Sponsor or an Investigator may take in order to protect the subjects of a trial against any immediate hazard to their health or safety. Upon implementation of an USM by an Investigator, the Sponsor must be notified immediately and details of the USM given. The Sponsor must immediately inform the REC by telephone, and then within 3 days in writing, setting out the reason/s for the safety measure and the plan for future action in accordance with the Sponsor’s standard operating procedures.

## CLINICAL INVESTIGATION MANAGEMENT

### Trial Steering Committee (TSC)

A TSC will be convened to provide independent review and overall supervision of the clinical investigation. The role of the TSC is to monitor progress and supervise the investigation to ensure it is conducted to high standards in accordance with the CIP, the principals of GCP, relevant regulations and guidelines with regards to participant safety.

A steering committee charter will be drawn up detailing the roles and responsibilities of the committee members.

### Data & Safety Monitoring Committee (DSMC)

A DSMC will be established to provide independent review. The purpose of the DSMC will be to monitor efficacy, safety and compliance data. The study DSMC will adopt a DAMOCLES charter which defines its terms of reference and operation in relation to oversight of the trial. In strict confidence, the DSMC will review accruing data, summaries of the data presented by treatment group, and will assess the screening algorithm against the eligibility criteria; together with any other analyses that the committee may request.

A DSMC charter will be drawn up detailing the roles and responsibilities of the committee members.

The DSMC will convene 6 weeks after the initial safety phase. Following a review of the safety data to date, they will advise on whether the RCT can commence.

#### PATIENT AND PUBLIC INVOLVEMENT (PPI)

PPI contributors have been, and will continue to be, actively involved in areas of the research such as:

Design of the research

Management and oversight of the research.

Development and review of patient information resources. Reporting of the research.

Dissemination of research findings.

## ETHICAL AND REGULATORY CONSIDERATIONS

### Declaration of Helsinki and Guidelines for Good Clinical Practice

The trial will be conducted in compliance with the approved clinical investigation plan and standard operating procedures (SOPs), the Declaration of Helsinki, the principles of Good Clinical Practice (GCP as stated in ISO 14155), the UK Data Protection Act and all other applicable regulatory and governance frameworks including the UK policy framework for health and social care research.

### Approvals

The CIP, Informed Consent Form, Participant Information Sheet and any proposed advertising material will be submitted to an appropriate Research Ethics Committee (REC), HRA (where required), regulatory authorities (MHRA in the UK), and host institution(s) for written approval. The investigator will submit and obtain approval from the above parties for all substantial amendments to the original approved documents.

The clinical investigation shall not begin until all the necessary regulatory approvals are in place.

If necessary, any additional requirements imposed by the REC or regulatory authority will be followed.

### Amendments to the CIP

Amendments to the CIP will be submitted to the sponsor for review and approval before being submitted to the REC and HRA for ethical approval and the MHRA as the relevant Competent Authority (CA) for the clinical investigation.

Before implementation of an amendment documented evidence of a favourable opinion will have been received from the relevant Research Ethics Committee (REC) and a notice of amendment acceptable from the MHRA.

A new (approved) version of the CIP will be distributed to participating centres with evidence of approval.

Original documentation relating to the amendment will be filed in the TMF.

### Reporting

The CI shall submit once a year throughout the clinical investigation, or on request, an Annual Progress Report to the REC, HRA (where required), host organisation and sponsor. In addition, an End of Trial notification and final report will be submitted to the MHRA, the REC, host organisation and sponsor.

### Participant Confidentiality

Staff will ensure that the participants’ anonymity is maintained. The clinical investigation will comply with the current UK Data Protection Act 2018 and UKGDPR which requires data to be anonymised as soon as it is practical to do so. All documents will be stored securely and only accessible by clinical investigation staff and authorised personnel.

Consent will be obtained from patients for personal information such as; name, address, email and telephone number to facilitate questionnaire follow-up.

Consent will be obtained for images and radiological reports to be sent to central study team with identifiable data such as the name, date of birth, NHS number and study number of the participant as this needs to be linked up with NHS patient records to link with the personalised implant at the time of surgery.

Consent will also be obtained for images and radiological reports to be sent to the device manufacturer, containing NHS number and unique study number.

### Expenses and Benefits

Reasonable travel expenses for any visits additional to normal care will be reimbursed on production of receipts, or a mileage allowance provided as appropriate.

### Training

All staff involved in the PASHiOn trial must be appropriately trained, including GCP training, to ensure that they are suitably experienced to perform the specific tasks that they are being asked to undertake as part of the trial. Evidence of GCP training for central study staff will be saved on a central system. Copies of GCP training certificates will be held centrally in the eTMF for each site investigator (PI and Co-Is). Evidence of GCP training for all other site staff members will be stored locally in the ISF.

### Other Ethical Considerations

The investigation will undergo review by the Medicines and Healthcare products Regulatory Agency (MHRA) and an NHS Research Ethics Committee to ensure the investigational device is safe for use in participants of the clinical investigation.

No treatment will be withheld. Patients will still receive standard care for their condition.

## FINANCING AND INSURANCE

### Funding

The PASHiOn clinical investigation is funded by Versus Arthritis. The Cardiff biomechanics sub-study is funded by ACCELERATE ‘Welsh Health Innovation Technology Accelerator’ through the ACCELERATE partner, Clinical Innovation Accelerator (Cardiff University). ACCELERATE is part funded by the European Regional Development Fund.

### Insurance

The University has a specialist insurance policy in place which would operate in the event of any participant suffering harm as a result of their involvement in the research NHS indemnity operates in respect of the clinical treatment that is provided.

### PUBLICATION POLICY

The results manuscript will be submitted for publication in peer reviewed journals, with the aim to publish in high-impact journals as open-access publications. Shorter articles on different aspects of the findings of the clinical investigation may also be published.

The investigators will be involved in reviewing drafts of the manuscripts, abstracts, press releases and any other publications arising from the clinical investigation. Authors will acknowledge that the clinical investigation was funded by Versus Arthritis. Authorship will be determined in accordance with the ICMJE guidelines and other contributors will be acknowledged. All members of the PASHiOn Clinical Investigation Group will be submitted to be listed and citable in PubMed.

## Data Availability

This is the study protocol for the PASHiOn trial and as such data availability is not relevant.

## APPENDICES

## Appendix A: Follow up schedule and strategy to account for delays in surgery and aligning post randomisation assessment with routine clinical follow up

| Type | Time zero | Randomisati<br>on to<br>Surgery<br>(months) | Primary<br>outcome<br>assessment<br>point (months<br>from surgery) | Prim follow<br>FU time<br>from<br>randomisati<br>on | Comments | Action |
| --- | --- | --- | --- | --- | --- | --- |
| A | Randm | 1 | 6 | 7 | Early surg. Unlikely – but can be accepted. Data point 2/12 earlier from rand | NONE |
| A | Randm | 2 | 6 | 8 | Early surg. Unlikely – but can be accepted. Data point 1/12 earlier from rand | NONE |
| P | Randm | 3 | 6 | 9 | IDEAL – no delay and 6/12 clinical and 9/12 from randomised data point aligned. | NONE |
| B | Randm | 4 | 5 | 9 | Allows for short delay. Acceptable. Time from rand within protocol. | bring patient in 1/12 earlier for clin assess. |
| B | Randm | 5 | 4 | 9 | Allows for short delay. Acceptable. Time from rand within protocol. | bring patient in 2/12 earlier for clin assess. |
| C | Randm | 6 | 4 | 10 | Allows for longer delay. Data still OK but data point outside ideal protocol (prolonged) | bring patient in 2/12 earlier for clin assess. |
| C | Randm | 7 | 4 | 11 | Allows for longer delay. Data still OK but data point outside ideal protocol (prolonged) | bring patient in 2/12 earlier for clin assess. |
| C | Randm | 8 | 4 | 12 | Allows for longer delay. Data still OK but data point outside ideal protocol (prolonged) | bring patient in 2/12 earlier for clin assess. |
| C | Randm | 9 | 4 | 13 | Allows for longer delay. Data still OK but data point outside ideal protocol (prolonged) | bring patient in 2/12 earlier for clin assess. |
| C | Randm | 10 | 4 | 14 | Allows for longer delay. Data still OK but data point outside ideal protocol (prolonged) | bring patient in 2/12 earlier for clin assess. |
| C | Randm | 11 | 4 | 15 | Allows for longer delay. Data still OK but data point outside ideal protocol (prolonged) | bring patient in 2/12 earlier for clin assess. |

## Appendix B: Schedule of Assessments

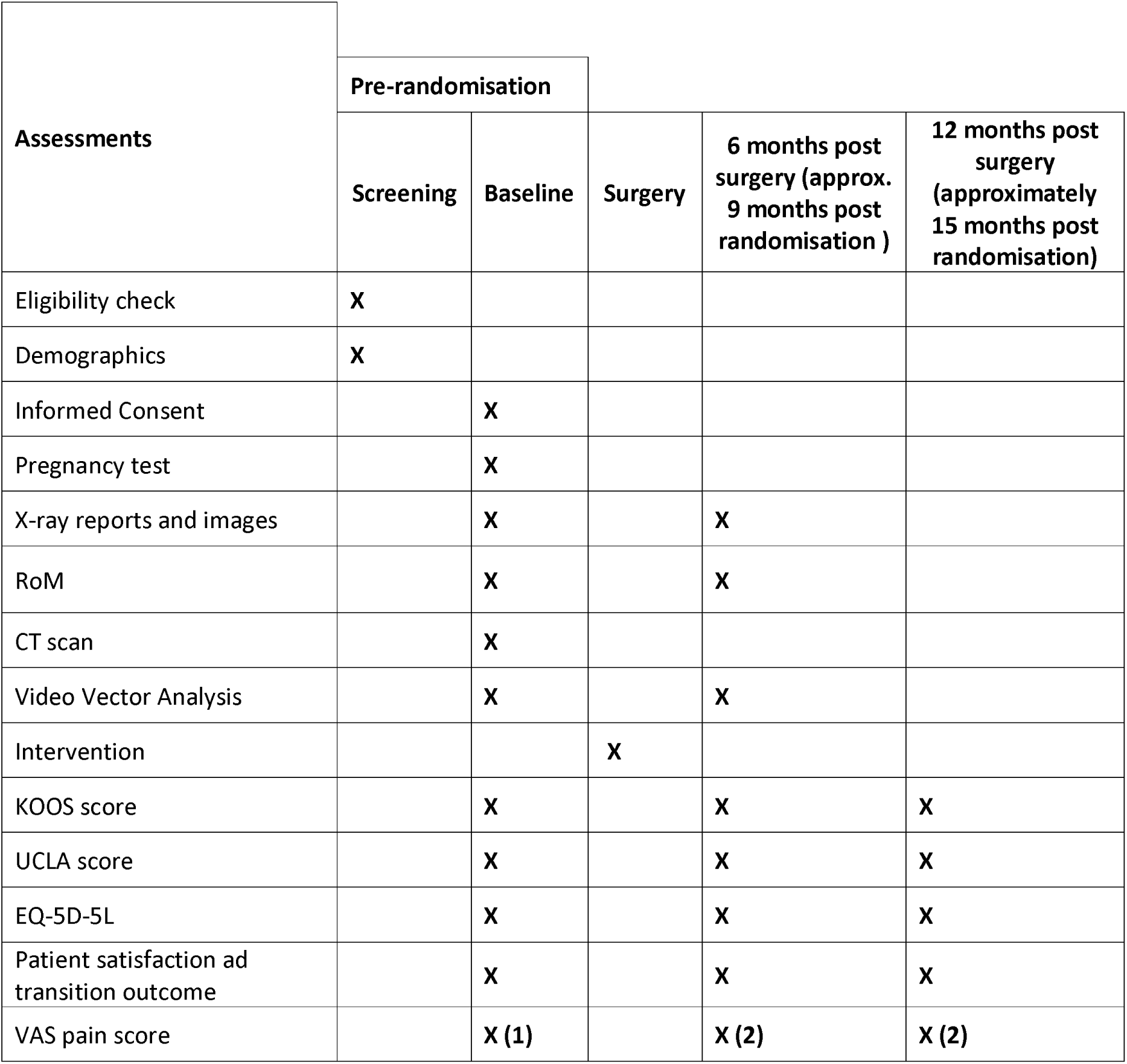

## Appendix C: Participant Flow for RCT (Phase 2) Note: Patients in the safety phase (Phase 1) will follow the same participant flow. They will be recruited and assessed in an identical way to Phase 2, but without randomisation

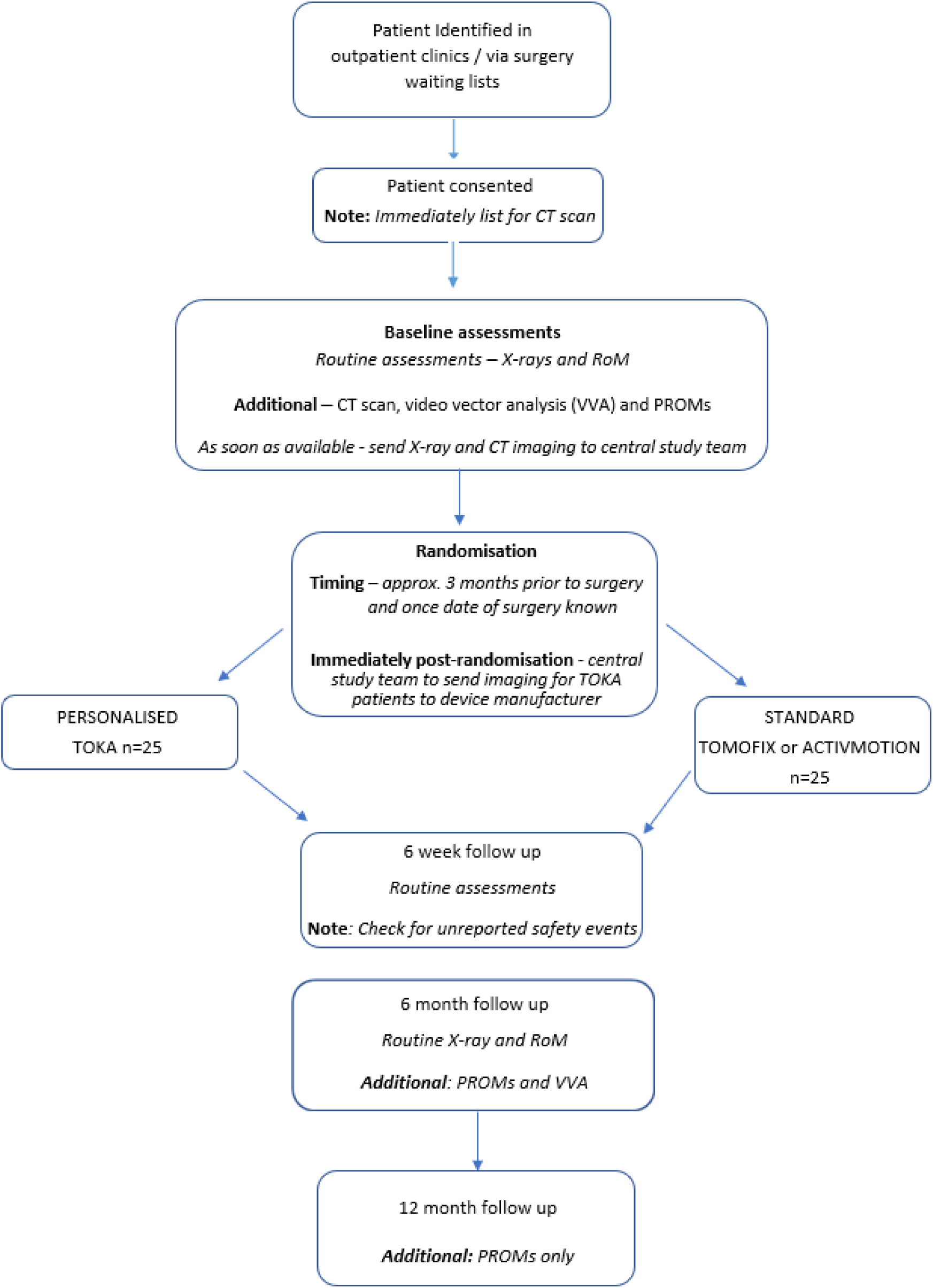

## Appendix D: Decision tree for adverse event reporting – Medical Devices 86565123-file00.docx

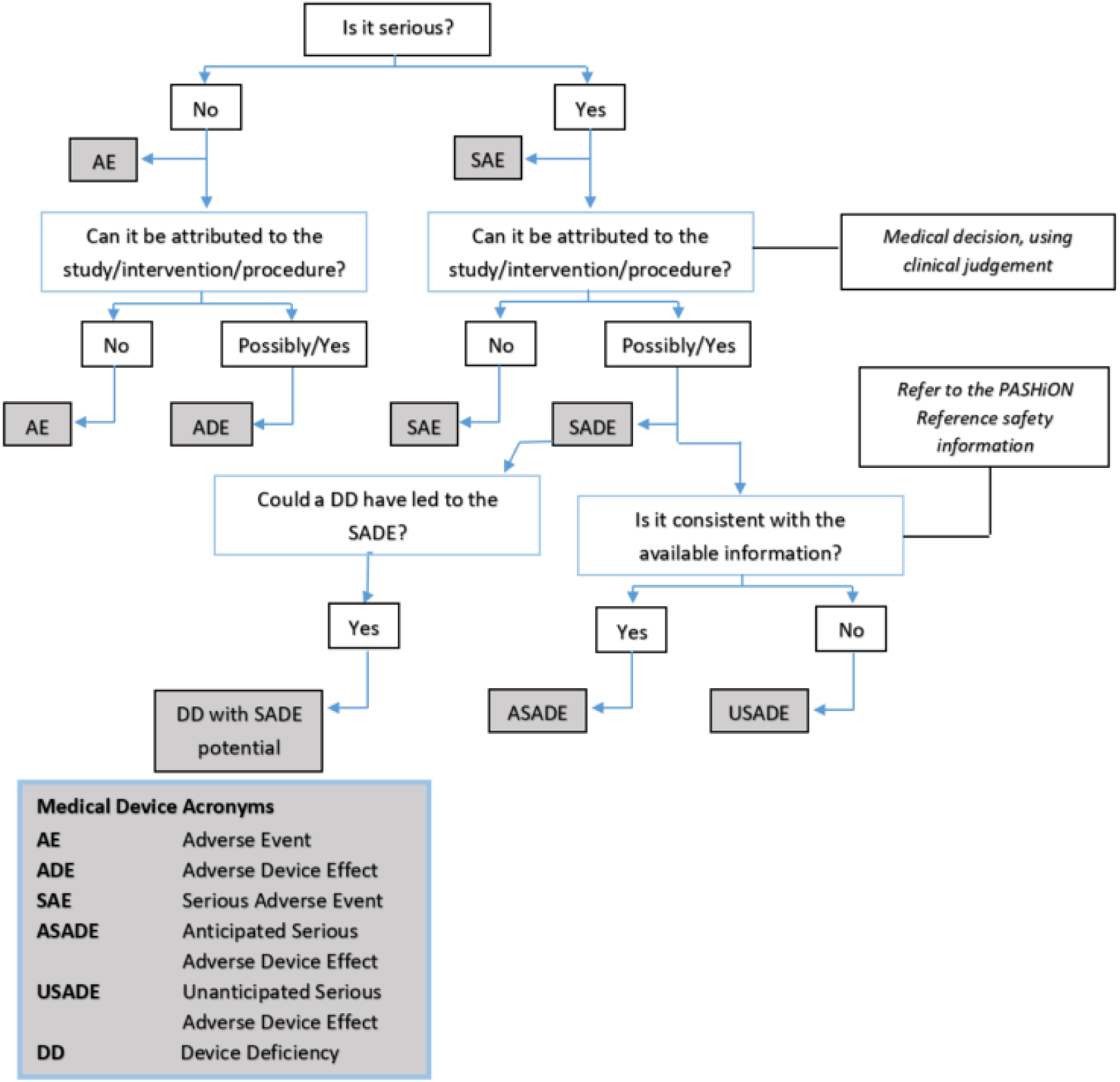

## Appendix E: Statistical Analysis Plan

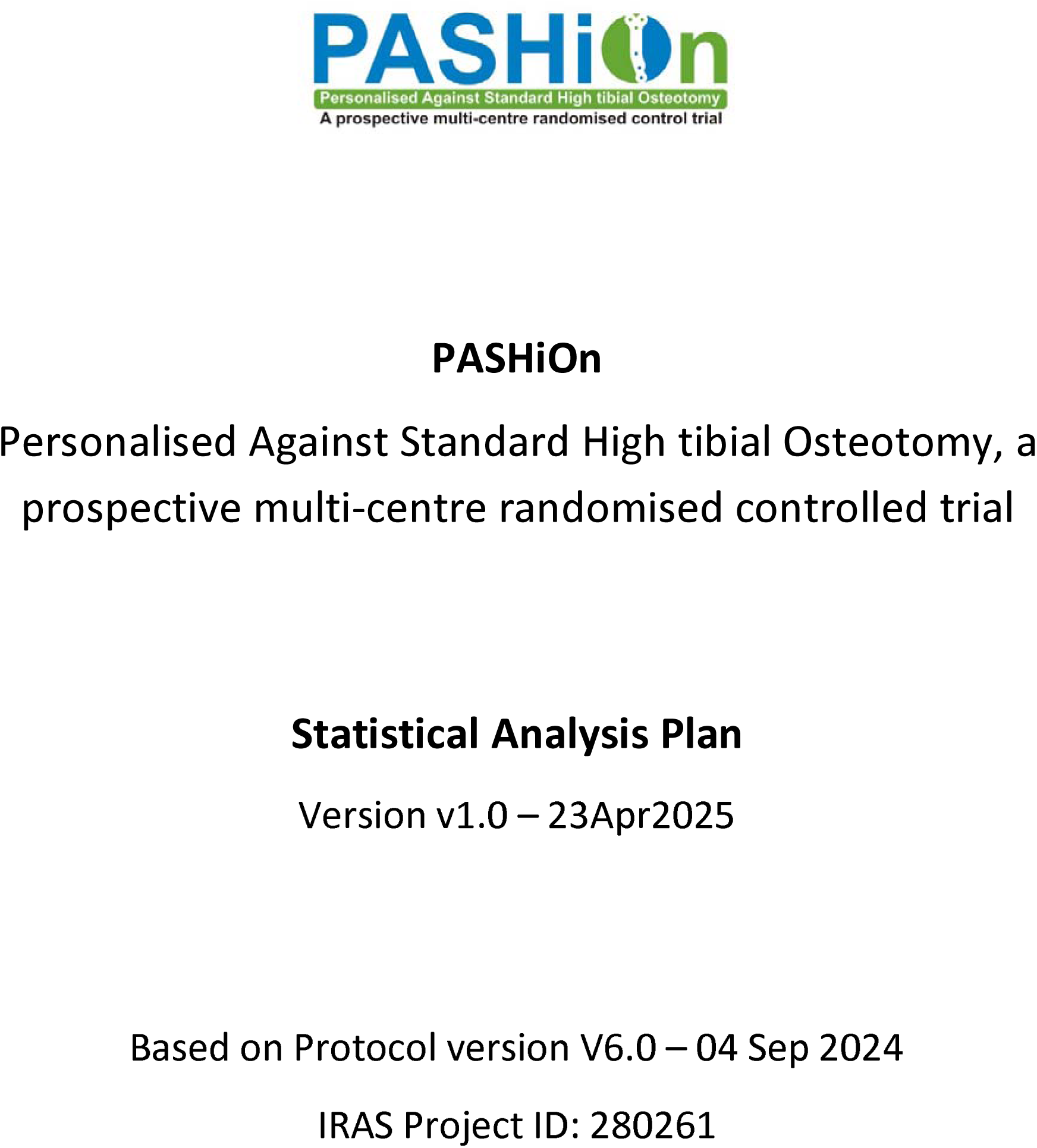
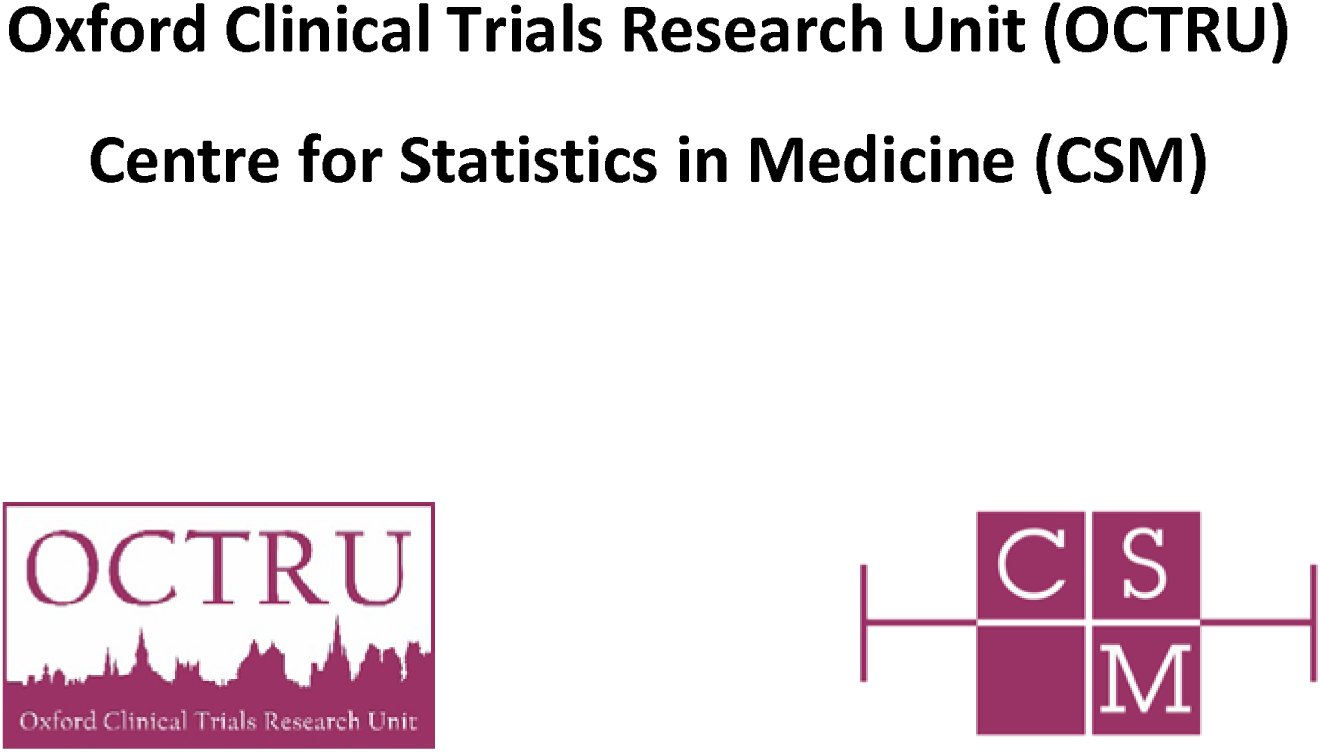

PASHiOn

Personalised Against Standard High tibial Osteotomy, a prospective multi-centre randomised controlled trial

Statistical Analysis Plan

Version v1.0 – 23Apr2025

Based on Protocol version V6.0 – 04 Sep 2024 IRAS Project ID: 280261

Oxford Clinical Trials Research Unit (OCTRU) Centre for Statistics in Medicine (CSM)

## 1. Introduction

This document details the proposed data presentation and analysis for the main paper(s) and final study reports from the Versus Arthritis funded prospective multi-centre randomised controlled trial comparing personalised against standard high tibial osteotomy (PASHiOn). The results reported in these papers should follow the strategy set out here. Subsequent analyses of a more exploratory nature or of extended study follow-up will not be bound by this strategy unless explicitly stated to be covered, though they are expected to follow the broad principles laid down here. The principles are not intended to curtail exploratory analysis (for example, to decide cut-points for categorisation of continuous variables), nor to prohibit accepted practices (for example, data transformation prior to analysis), but they are intended to establish the rules that will be followed, as closely as possible, when analysing and reporting the trial. This document follows published guidelines regarding the content of statistical analysis plans for clinical trial (1).

The analysis strategy will be available on request when the principal papers are submitted for publication in a journal. Suggestions for subsequent analyses by journal editors or referees, will be considered carefully, and carried out as far as possible in line with the principles of this analysis strategy. If reported, the analyses will be marked as post-hoc; the source of the suggestion will be acknowledged, and the reader will be advised to rely primarily on the pre-specified analysis for the interpretation of the results.

Any deviations from the statistical analysis plan will be described and justified in the final report of the trial. The analysis should be carried out by an identified, appropriately qualified and experienced statistician, who should ensure the integrity of the data during their processing. Examples of such procedures include quality control and evaluation procedures.

### 1.1 Key personnel

List of key people involved in the preparing, reviewing and approving the SAP and subsequent reports.

Author:

Alexander Thomas

Reviewers: David Smith David Beard Alisdair MacLeod

Approvers: Jonathan Cook Richie Gill

### 1.2 Changes from previous version of SAP

A summary of key changes from earlier versions of SAP, with relevance to protocol changes that have an impact on the design, definition, sample size, data quality/collection and analysis of the outcomes will be provided. Include protocol version number and date.

| Version number<br>Issue date | Author<br>of this<br>issue | Protocol Version & Issue date | Significant changes from previous<br>version together with reasons |
| --- | --- | --- | --- |
| V1.0_DDMonYYYY |  | V6.0 04 Sep 2024 | Not applicable as this is the 1<sup>st</sup> issue |

## 2. Background and Objectives

Up to 28% of the UK population over 40 have knee pain, with half of these people having radiographic osteoarthritis. This leads to a high demand for knee replacement surgeries. It is important to recognise that one third of knee replacements are now performed in patients younger than 65 indicating the disease burden and need for treatment in a younger group of osteoarthritis sufferers. The current treatment choice is a total knee replacement; however, it is not recommended for the earlier stages of knee osteoarthritis and it does not last as long in younger patients. Patients aged 64 years or less at time of primary surgery have double the revision rate of those aged between 65 and 74.

An alternative to knee replacement is high tibial osteotomy, commonly referred to as HTO. In HTO, the native joint is preserved by realigning the tibia to off-load the worn areas of the knee. A well performed HTO can delay the need for knee replacement by 10 years. There are barriers in place preventing wide adoption of HTO – including the challenging operative technique. The gold standard for assessing lower limb alignment in the coronal plane is to measure the hip-knee-ankle angle on full wight bearing x-rays. This measure forms our primary endpoint as we will analyse the absolute difference between planned and achieved coronal plane correction.

A list of objectives (as documented in the protocol) is given in Table 1Table 1 along with the outcome measures that will be used to answer the objective.

**Table 1.**
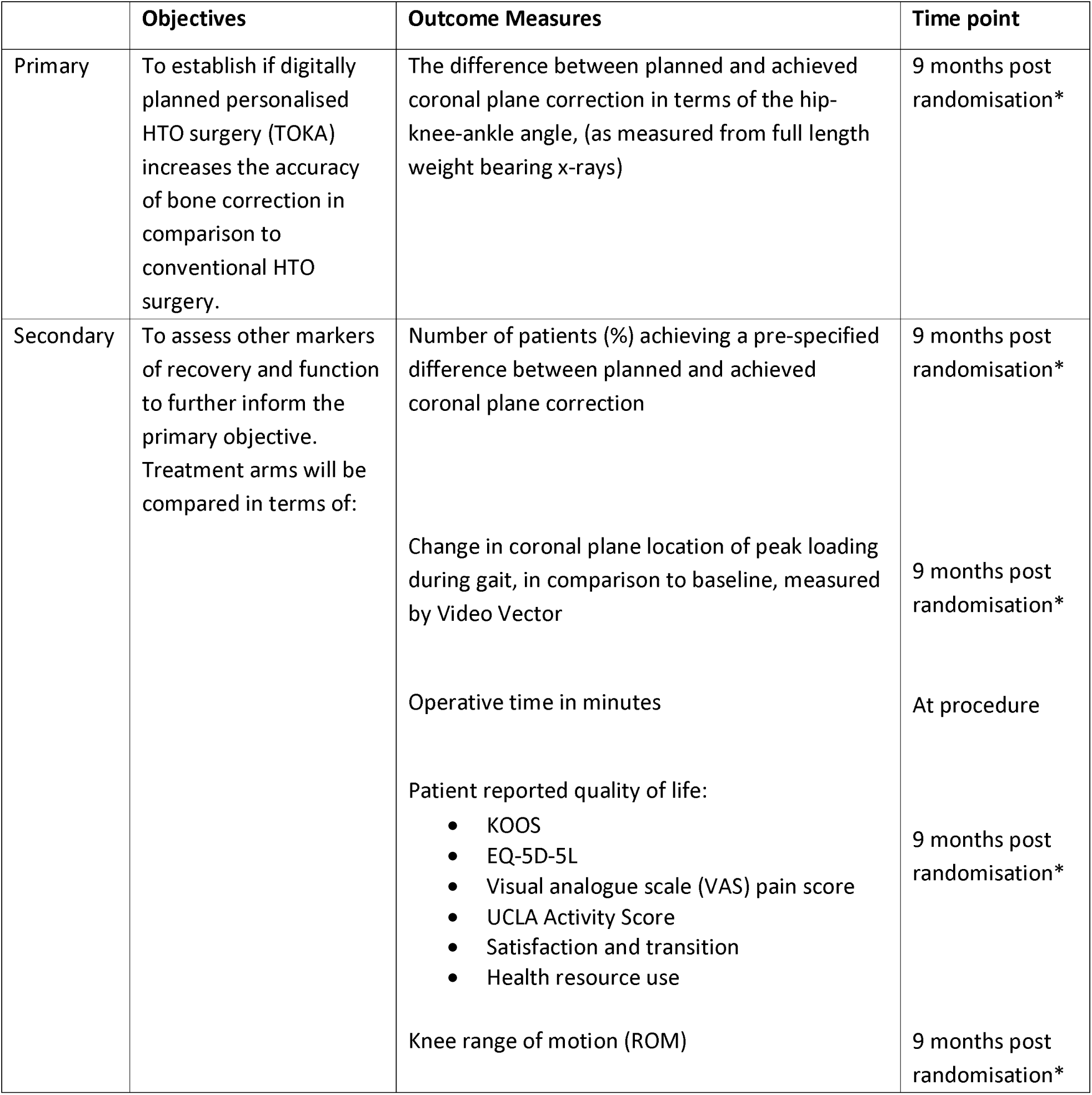
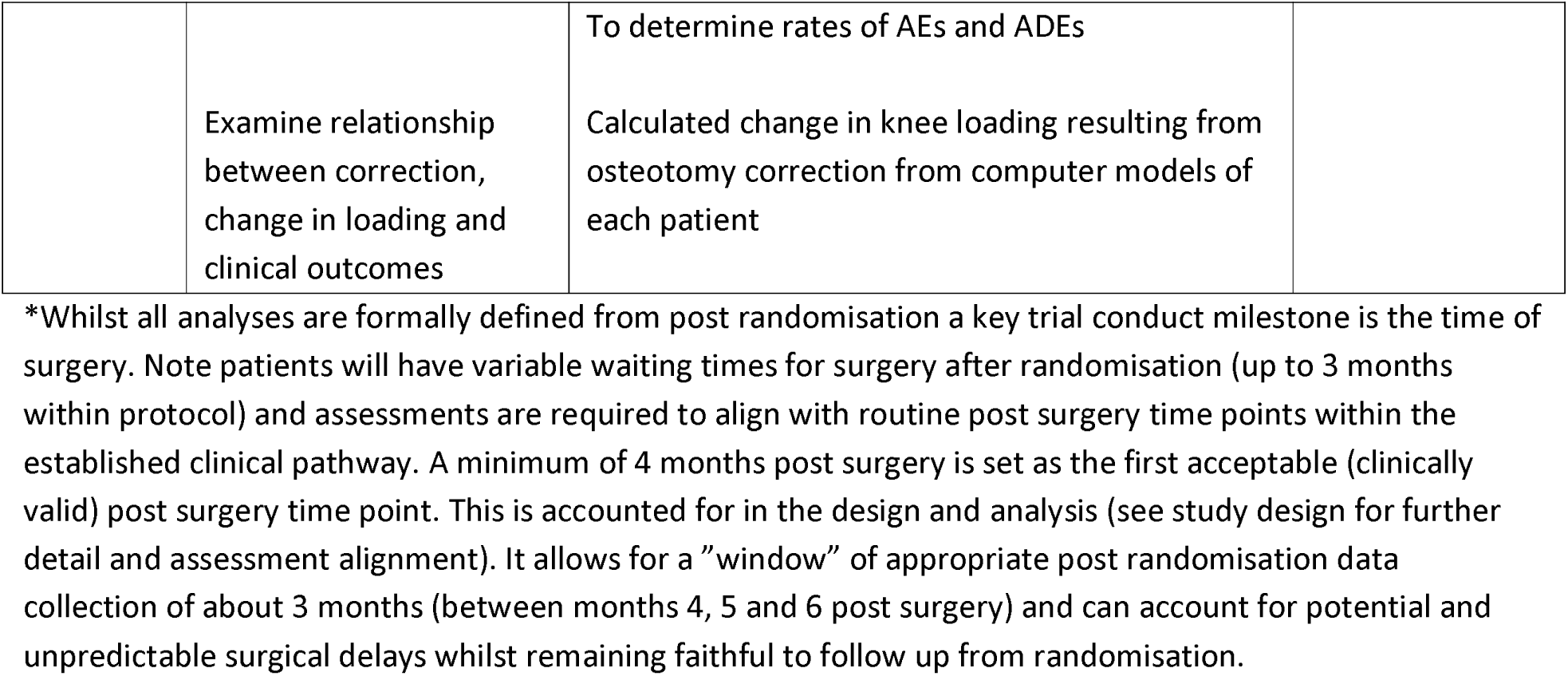
List of objectives, outcome measures and the time the measure will be taken.

|  | Objectives | Outcome Measures | Time point |
| --- | --- | --- | --- |
| Primary | To establish if digitally planned personalised HTO surgery (TOKA) increases the accuracy of bone correction in comparison to conventional HTO surgery. | The difference between planned and achieved coronal plane correction in terms of the hip-knee-ankle angle, (as measured from full length weight bearing x-rays) | 9 months post randomisation* |
| Secondary | To assess other markers of recovery and function to further inform the primary objective. Treatment arms will be compared in terms of: | Number of patients (%) achieving a pre-specified difference between planned and achieved coronal plane correction | 9 months post randomisation* |
|  |  | Change in coronal plane location of peak loading during gait, in comparison to baseline, measured by Video Vector | 9 months post randomisation* |
|  |  | Operative time in minutes | At procedure |
|  |  | Patient reported quality of life:<br>• KOOS<br>• EQ-5D-5L<br>• Visual analogue scale (VAS) pain score<br>• UCLA Activity Score<br>• Satisfaction and transition<br>• Health resource use | 9 months post randomisation* |
|  |  | Knee range of motion (ROM) | 9 months post randomisation* |

|  |  |  |
| --- | --- | --- |
|  | Examine relationship between correction, change in loading and clinical outcomes | To determine rates of AEs and ADEs<br>Calculated change in knee loading resulting from osteotomy correction from computer models of each patient |

\*Whilst all analyses are formally defined from post randomisation a key trial conduct milestone is the time of surgery. Note patients will have variable waiting times for surgery after randomisation (up to 3 months within protocol) and assessments are required to align with routine post surgery time points within the established clinical pathway. A minimum of 4 months post surgery is set as the first acceptable (clinically valid) post surgery time point. This is accounted for in the design and analysis (see study design for further detail and assessment alignment). It allows for a "window" of appropriate post randomisation data collection of about 3 months (between months 4, 5 and 6 post surgery) and can account for potential and unpredictable surgical delays whilst remaining faithful to follow up from randomisation.

**Table 2:**
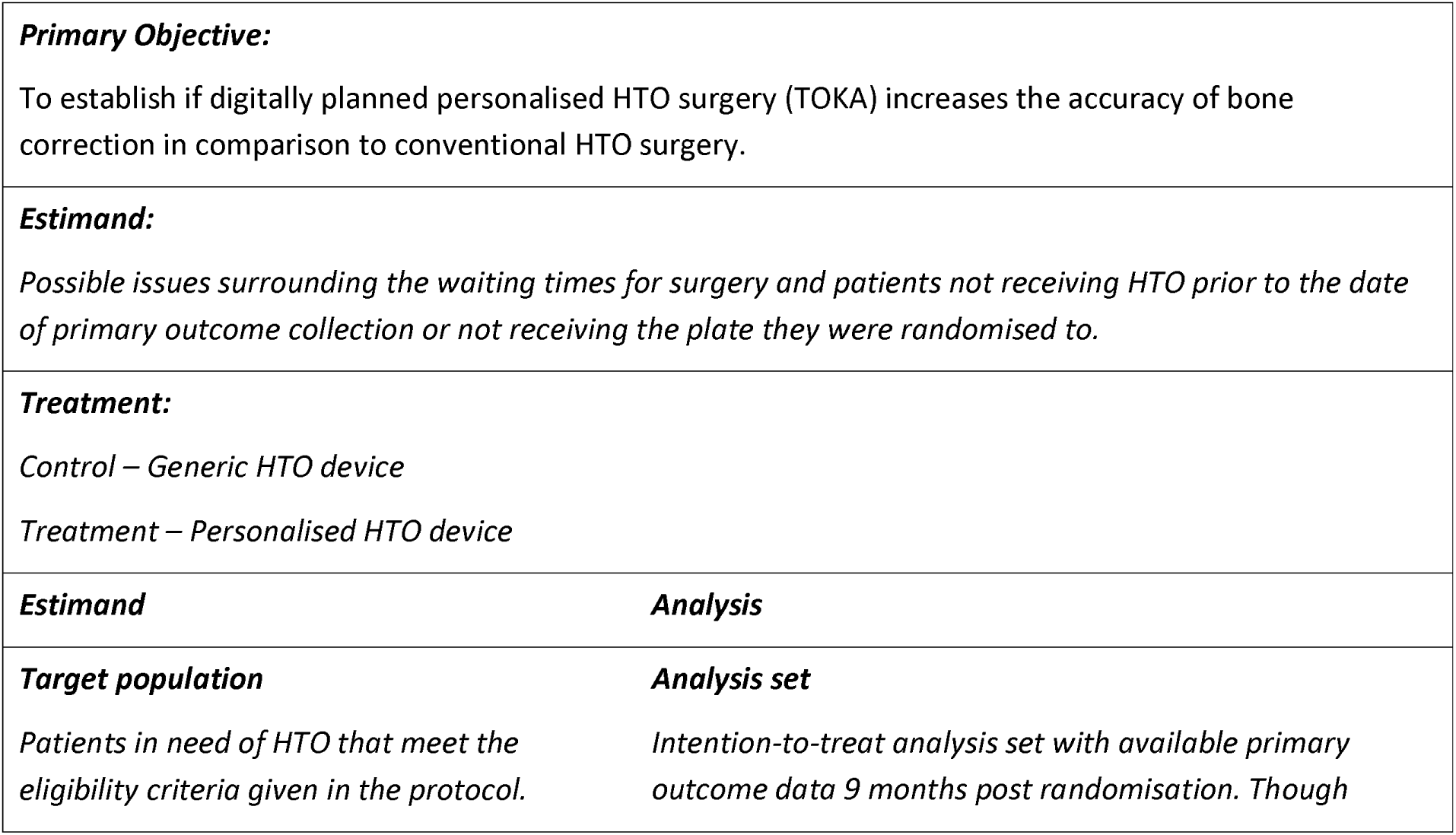
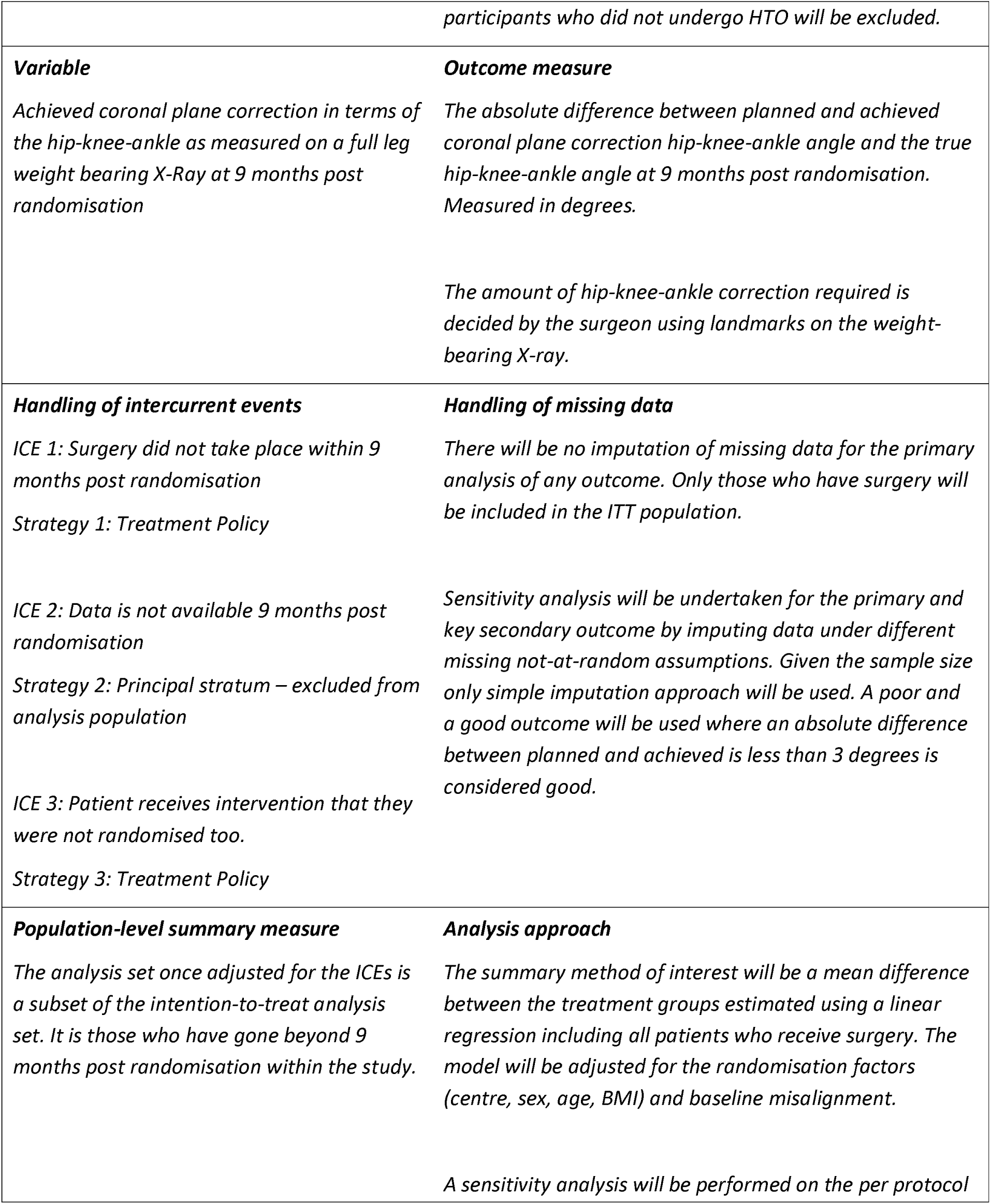
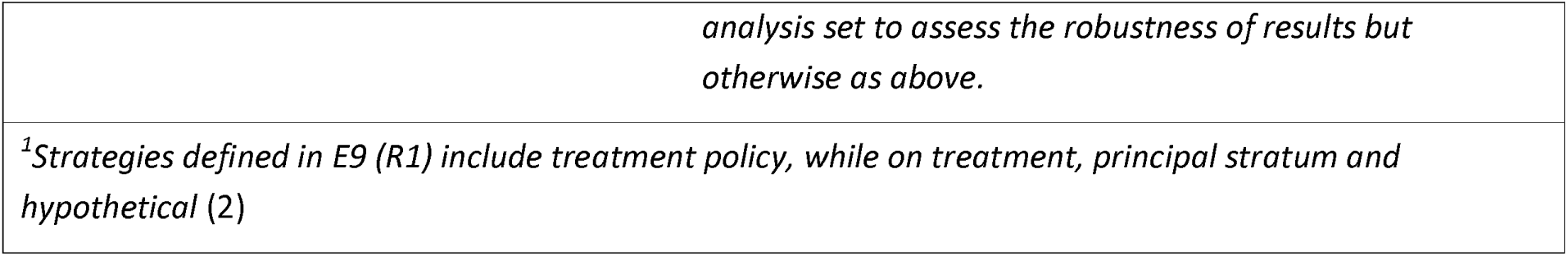
Estimand-to-analysis table.

## 3. Study Methods

### 3.1 Trial Design/framework

PASHiOn is a multi-centre, blinded, superiority two arm, parallel group design randomised controlled trial of personalised versus conventional HTO surgery. Patients will be randomised in a 1:1 ratio. Patients and primary outcome assessors will be blinded to the allocation.

### 3.2 Embedded within the trial is a non-randomised pre-trial technology check and safety assessment, this is termed the Initial Safety Phase (Phase 1). Randomisation and Blinding

The patients within Phase 1, are not randomised as they all received HTO for initial safety checks to be undertaken. For this reason, there was also no blinding within Phase 1.

Randomisation to Phase 2 will be 1:1 by minimisation, using the following factors: trial centre, sex, age and BMI. We have chosen to separate patients younger than 50 years and 50 years and older, and patients with a BMI lower than 30 and those with a BMI of 30 or greater, as stratification cut-off points.

The minimisation algorithm will include a random element to reduce predictability. A small number of participants will be randomised using a simple randomisation schedule generated in advance to patient randomisation.

All patients and outcome assessors will be blinded to the treatment allocation.

### 3.3 Sample Size

The sample size was initially planned to include 93 patients (inclusive of 5 participants in Phase 1 and 88 in Phase 2), this was recalculated and the target sample size was reduced to 50. The 50 will be achieved over 2 stages: Phase 1 – initial safety phase (5 participants) and Phase 2 – RCT (45 participants).

- Phase 1 – Initial safety phase (5 participants)
- Phase 2 – RCT (45 participants)

This sample size calculation was based upon the primary outcome of value of absolute difference between achieved and planned correction. A total sample size of 45 participants will be sufficient to detect a standardised difference of 2.6, using 80% power, a 5% significance level and allowing for 10% loss to follow up.

### 3.4 Statistical Interim Analysis, Data Review and Stopping guidelines

No interim analysis is planned, similarly no formal subgroup analysis is planned either. One single formal analysis will be conducted after the final follow-up assessment has been completed and sufficient time allowed for data collection and data cleaning.

The independent DSMC reviewed the safety data for the 5 patients recruited into Phase 1. Based upon this review, the DSMC confirmed the trial could proceed to Phase 2.

During Phase 2, the DSMC and TSC have evaluated the risk of the trial and appropriate actions have been taken where necessary. For more details, see the reports and the charter.

### 3.5 Timing of Final Analysis

Final analysis is expected to begin after the final follow-up assessment is completed, allowing for a reasonable time to complete data collection and data cleaning.

### 3.6 Blinded analysis

There is no plan to perform any formal blinded analysis.

### 3.7 Statistical Analysis Outline as presented in the protocol

A separate statistical analysis plan (SAP) with full details of all statistical analysis planned for the data in this study will finalised prior to any primary outcome analysis. The SAP will be reviewed and receive input from the Trial Steering Committee (TSC) and Data and Safety Monitoring Committee (DSMC). Any changes or deviations from the original SAP will be described and justified in the protocol, final report and/or publications, as appropriate. It is anticipated that all statistical analysis will be undertaken using Stata (StataCorp LP, www.stata.com) or another well-validated statistical package.

## 4. Statistical Principles

There is a single primary outcome, so there is no concern regarding multiple testing in regards to multiple primary outcomes. A 2-sided significance level of 0.05 will be used, with 95% confidence intervals reported. All secondary analyses will be considered as supporting the primary analysis and will also be analysed using a 2-sided significance level of 0.05 with 95% confidence intervals. Interim analyses of primary and secondary outcomes have not be carried out as was planned.

### 4.1 Definition of Analysis Populations

Populations for analysis are defined as follows:

- Intent-to-treat (ITT): all participants analysed in their randomised groups, regardless of actual treatment received as long as they received knee surgery of some kind.
- Per-Protocol (PP): participants who received the intervention as intended will be analysed according to the treatment they actually received. Participants will be excluded from the per-protocol population if:

- They did not receive the treatment to which they were randomised
- They did not provide sufficient follow-up data analysis
- They did not satisfy the eligibility criteria for the study
- They did not adhere exactly to the protocol

Their preliminary scans were at least 6 months prior to surgery

Their surgery was at least 6 months post-randomisation

Exact exclusion criteria for PP analysis will be confirmed prior to conducting the final analysis (in that the estimates will not be generated and no results will be shared before this).

## 5. Trial Population and Descriptive Analyses

Summary of flow of trial participants through the trial and baseline stratification, demographic and clinical characteristics of each group.

SAP Version No: 1.0 OCTRU-OT-

061_V6.0_15May2024

Date: 23 Apr 2025 Effective Date

29May2024

### 5.1 Representativeness of Study Sample and Patient Throughput

The flow of participants through the trial will be summarised as outlined in Figure 1. This shows the number of individuals screened, eligible, consented, randomised to each arm, receiving allocated treatment and included in primary analysis as recommended in the CONSORT guidelines (3). Reasons for ineligibility, withdrawals, and exclusions from primary analysis will be summarised.

**Figure 1.**
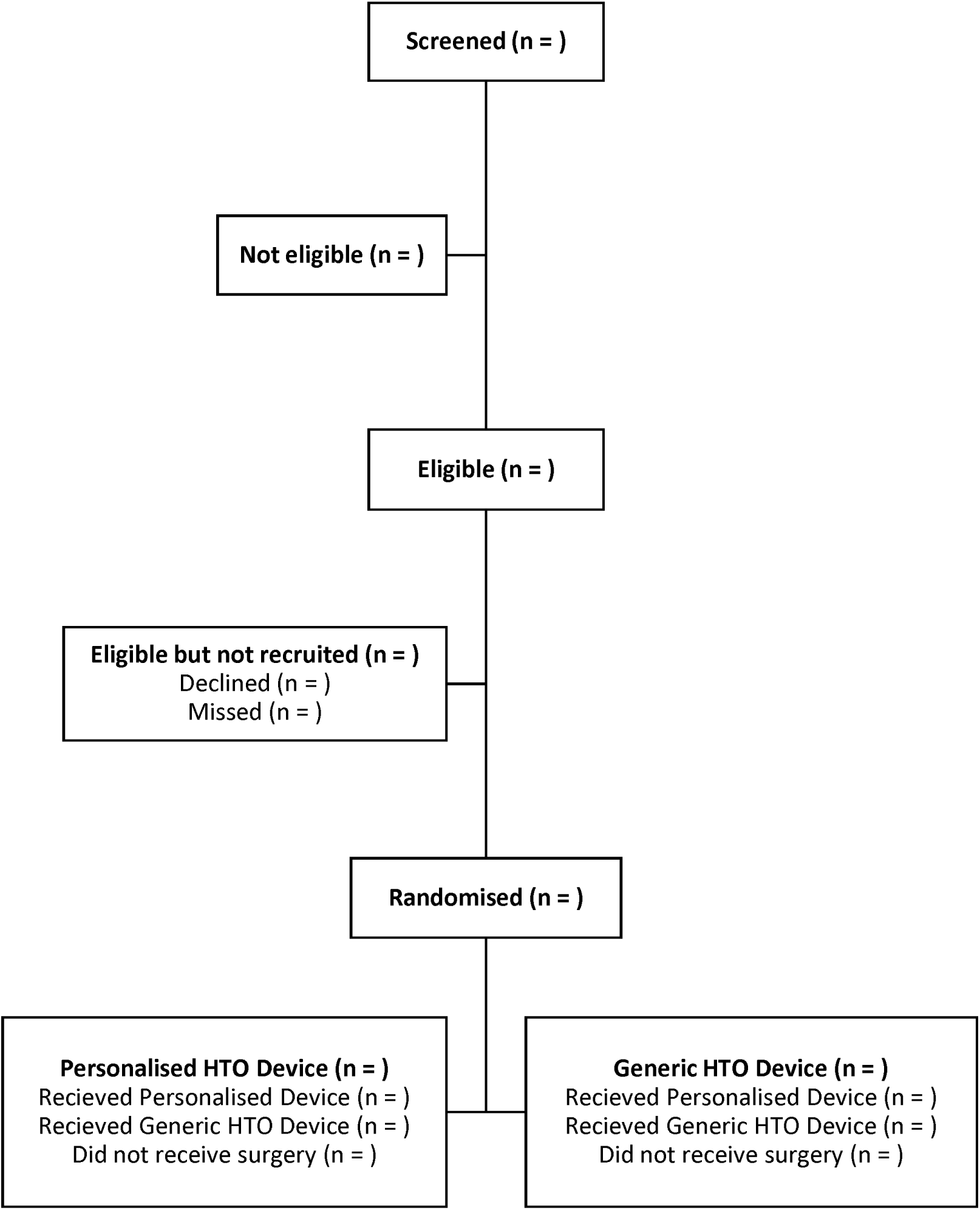
CONSORT Flow diagram.

### 5.2 Withdrawal from treatment and/or follow-up

The numbers and percentages of participants who are lost to follow-up or withdrew are reported (Table 3) by treatment allocation for each time point until the final time-point of 15 months post-randomisation.

Reasons for withdrawal will also be summarised by treatment allocation in Table 4.

**Table 3.** Details of withdrawal.

| Patient ID | Extent of withdrawal | Withdrawal Date (DDMonYYYY) | Withdrawal Reason | Last Completed Questionnaire | Data Available for Primary Outcome (Y/N) |
| --- | --- | --- | --- | --- | --- |
| ID 1 | Intervention only vs. follow-up |  |  |  |  |
| ID 2 |  |  |  |  |  |
| ID 3 |  |  |  |  |  |
| ... |  |  |  |  |  |

**Table 4.**
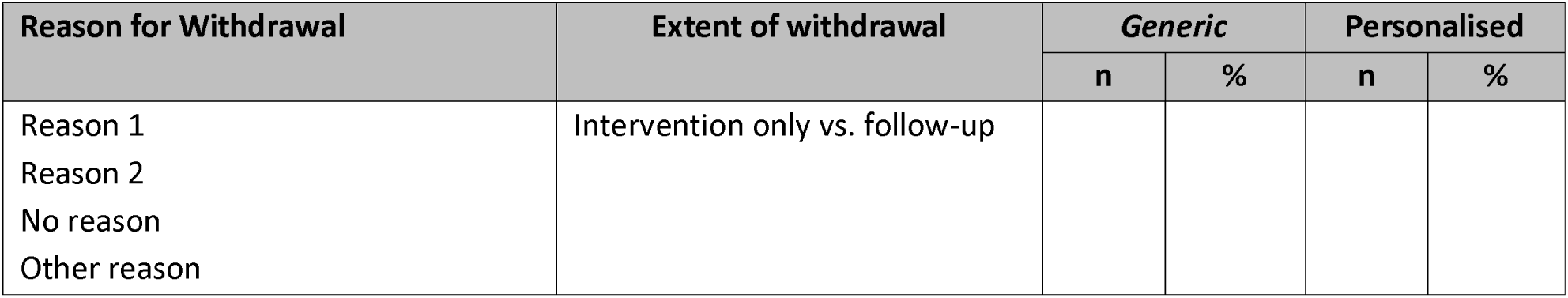
Summary of reasons for withdrawal by treatment arm.

### 5.3 Baseline Comparability of Randomised Groups

Baseline comparability of the randomised groups on the stratification factors (centre, sex, age, and BMI) will be presented in Table 5. Baseline comparability of the randomised groups on other factors will be presented (Table 6 and Table 7). Variables will be presented by numbers (with percentages) for binary and categorical variables. Means and standard deviations (SDs), or medians and interquartile ranges (IQRs) for continuous variables will be presented; there will be no tests of statistical significance nor confidence intervals for differences between randomised groups.

**Table 5.**
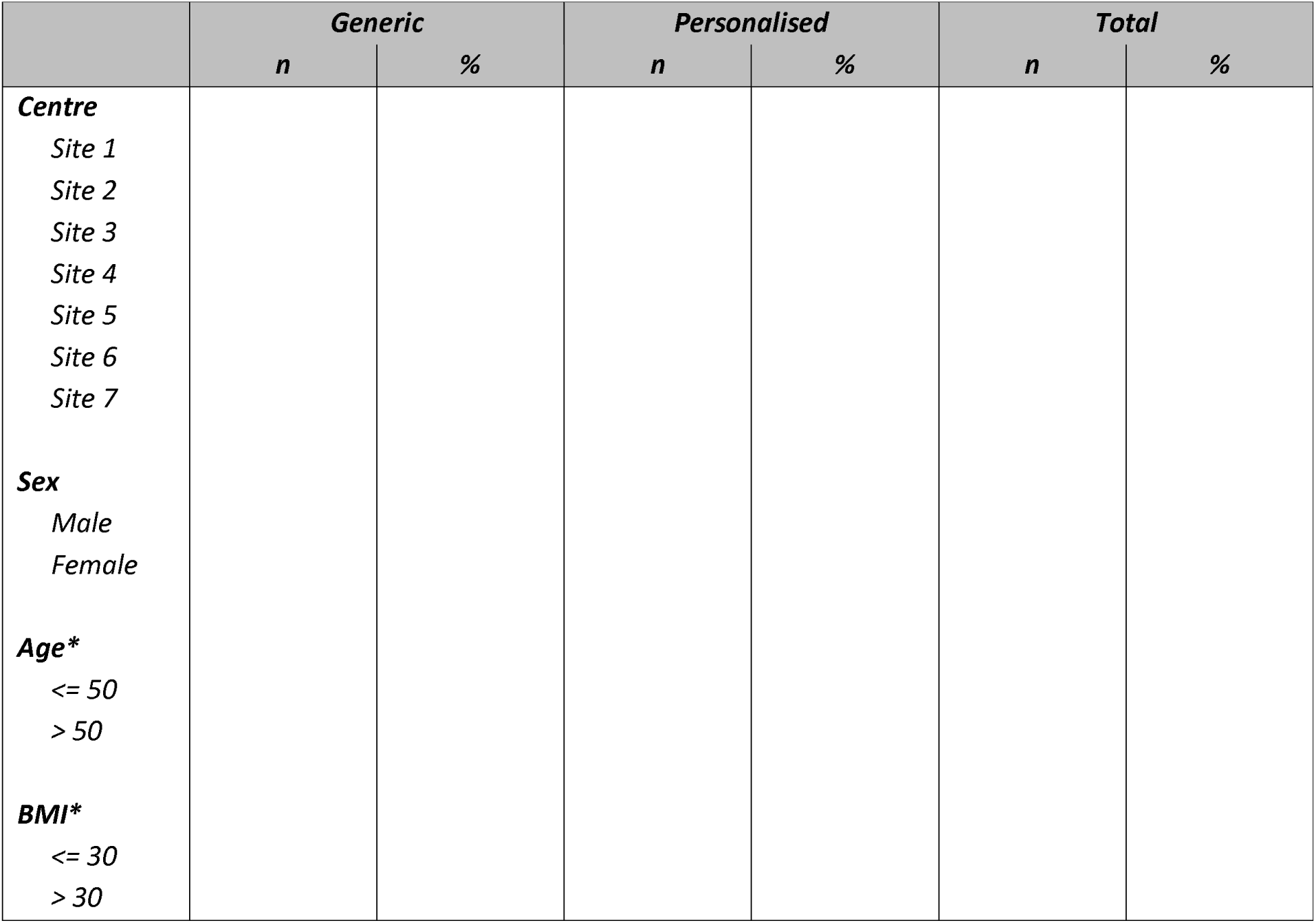
Stratification factors by treatment arm.

**Table 6.**
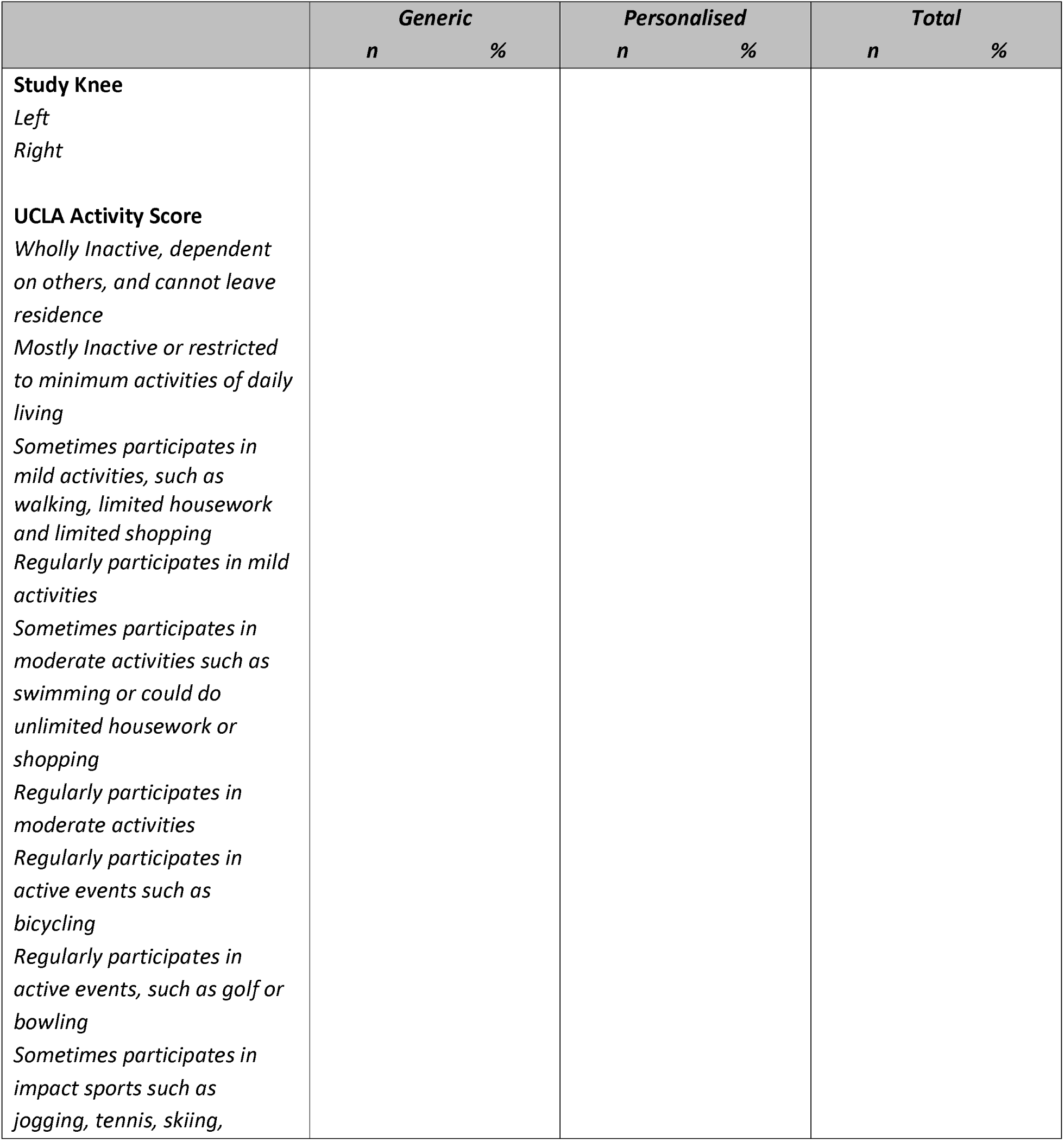
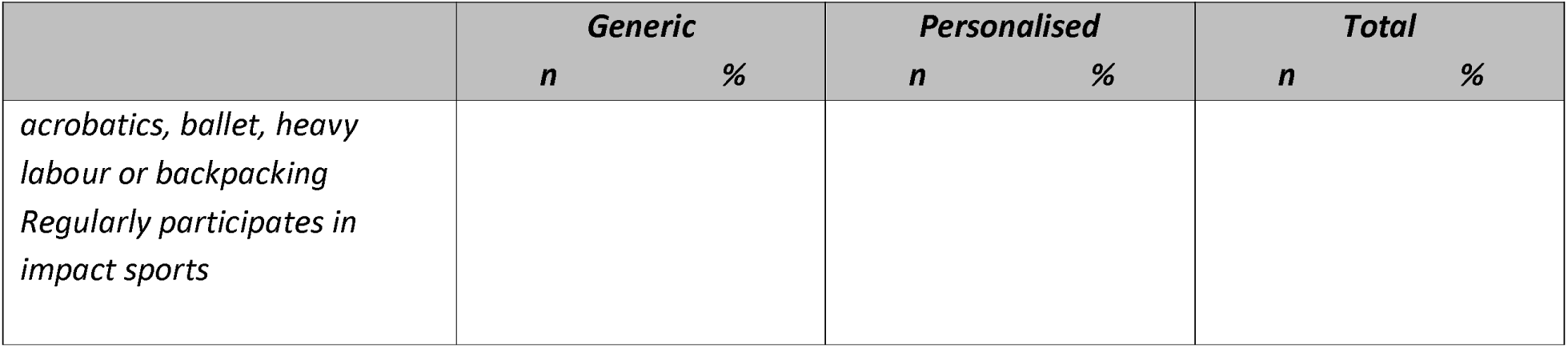
Categorical factors by treatment arm.

**Table 7.**
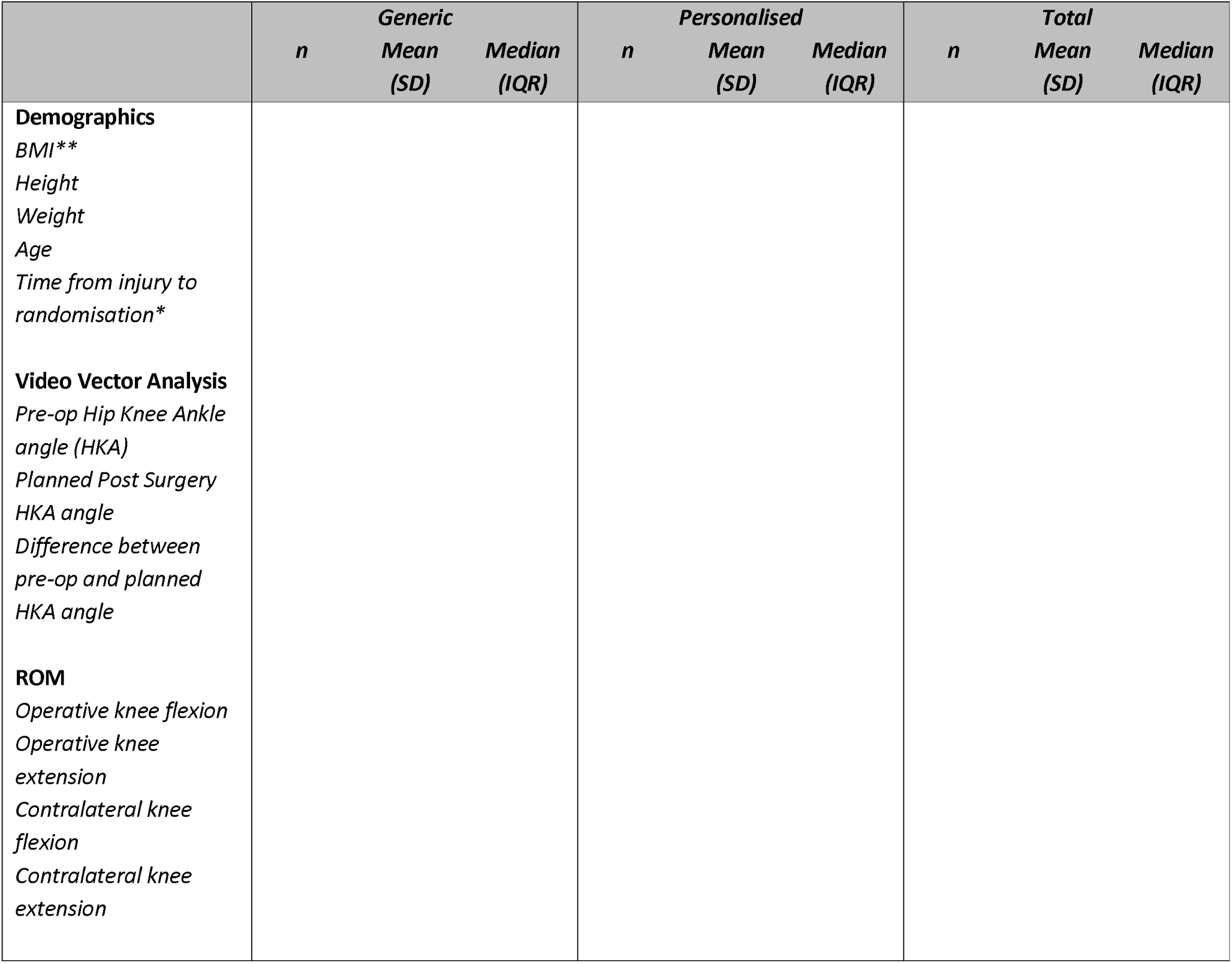
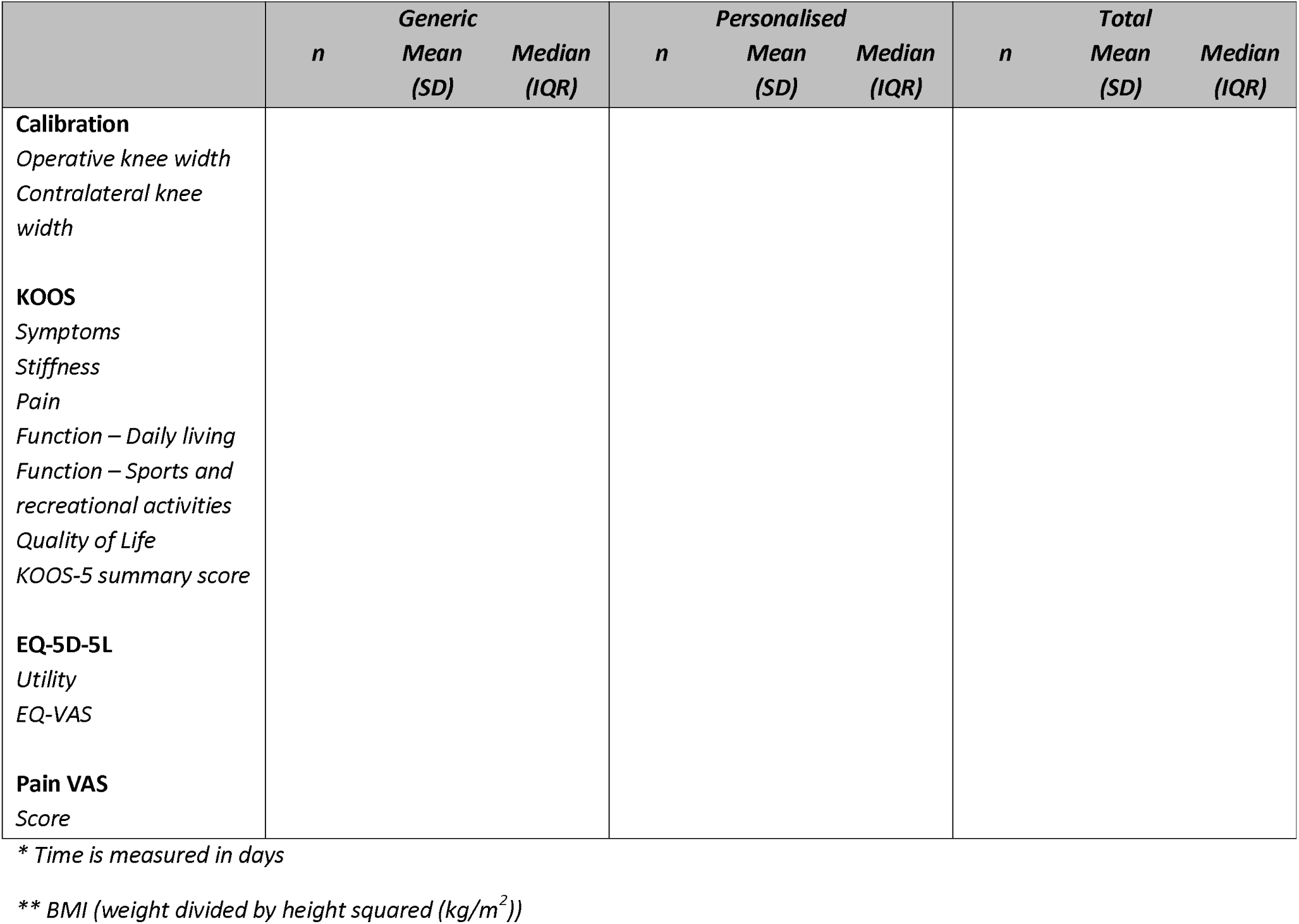
Continuous factors by treatment arm.

### 5.4 Unblinding

If patient unblinding is deemed necessary, it will be discussed on a case-by-case basis with the central study team and local PI. All unblinding will be at the discretion of the local investigators, when clinically indicated for the safety of the patient. There will be no formal analysis of the success of the blinding. All cases of treatment unblinding will be listed, together with who was unblinded, and reasons for unblinding summarised with numbers and percentages reported.

### 5.5 Treatment Compliance with Details of Interventions

Compliance with the interventions according to random allocation in this study will be presented as numbers and percentages of participants receiving their allocated treatment. The investigational device is a single-use implant, so the proportion of participants that received their allocated treatment defines treatment compliance. As such, participant compliance with the device is not relevant. If, for any reason, the randomised treatment (device) is not used this will be recorded and presented. Operation and theatre times will be summarised descriptively. Overall data per arm and per surgeon/site will be summarised as appropriate, including exploring any indication of learning.

### 5.6 Reliability

Sites will take the responsibility to double check and confirm the data collected. This includes registration data, the primary outcome (hip-knee-ankle angle measurement), and any withdrawal/death notifications. This data will be collected from trial sites directly and will be sent to the trial team on a regular basis.

It is worth noting that the target HKA alignment is predetermined for participants receiving the personalised HTO device in advance of surgery allowing for time to design the device. Conversely, participants receiving the generic HTO device have their target alignment determined on the date of surgery and completed in the CRF post-surgery. No methods are planned to counteract this potential bias in favour of the generic device.

## 6. Analysis

### 6.1 Outcome Definitions

#### Primary Outcome Definition

A full leg weight bearing X-Ray will be taken at baseline and at 9 months post randomisation. The images will be used to calculate the difference between planned and achieved coronal plane correction. Note for all measures 9 months post randomisation are intended to align with the 6 months post-surgery within the standard protocol (surgery within 3 months of randomisation and no delays). These time points can therefore be used interchangeably for the purposes of trial management.

The amount of Hip Knee Ankle (HKA) correction required is decided by the surgeon using landmarks on the weight-bearing X-ray. The Hip-Knee-Ankle (HKA) angle is defined as the lateral angle between two lines: one line from the centre of the femur head using Mose circles to the middle of the distance between the tibial spines, and a second line from the centre of the ankle to the centre of the tibial spines. An angle of more than 180 degrees denotes a varus alignment. The baseline HKA angle and the planned HKA correction angle is recorded pre-operatively. The HKA correction angle achieved is recorded post-operatively.

#### Secondary Outcome Definitions

- Number of patients achieving a pre-specified difference, 3 degrees, between planned and achieved coronal plane correction of the HKA. The HKA is calculated as per the primary outcome and converted into a binary variable difference (with 3 degrees) or not.
- Change in coronal plane location of peak loading during gait: Video Vector Analysis (VVA) will be used at baseline and at 6 months post-surgery. VVA simultaneously records ground reaction force data from a force plate and video data from a video camera focussed on the lower limb of a participant. This enables the magnitude, orientation, and location of the ground reaction force to be determined relative to the limb under investigation.
- Range of Motion (ROM): ROM flexion and extension will be measured (in degrees) at baseline and nine months post randomisation.
- Operative time in minutes: The difference in operative time and theatre time between the treatment arms. Operative time is defined as time from ‘knife to skin’ to time dressings have been applied; theatre time is defined as the time the participants enters the anaesthetic room to when they leave the operating theatre.
- KOOS: Knee injury and osteoarthritis Outcome score (KOOS) is a patient reported outcome measure derived from 5 subscales; symptoms (including stiffness), pain, function (daily living), function (sports and recreation activities) and quality of life with scores ranging from 0 – 100, a higher score indicating better health.

- `o` Scoring: Each subscale score is calculated separately, calculate mean score (x100) of individuals items of each subscale and divide by 4 (the highest possible score for a single question). Then take 100 – the score calculated. While there is no overall KOOS score the summary score using an average of the 5 domains has been suggested and used in previous clinical trials.
- `o` Missing data rules: If a mark is placed outside a box, the closest box is chosen. If two boxes are marked, that which indicates the more severe problem is chosen. As long as at least 50% of the subscale items are answered for each subscale, a mean score can be calculated. If more than 50% of the subscale items are omitted, the response is considered invalid and no subscale score should be calculated. For the subscale Pain, this means that 5 items must be answered; for Symptoms, 4 items; for ADL, 9 items; for Sport/Rec, 3 items; and for QOL, 2 items must be answered in order to calculate a subscale score. Subscale scores are independent and can be reported for any number of the individual subscales, i.e. if a particular subscale is not considered valid (for example, the subscale Sport/Rec 2 weeks after total knee replacement), the results from the other subscale can be reported at this time-point.
- EQ-5D-5L: To measure quality of life the EQ-5D-5L will be collected at baseline, 6 and 12 months. The EQ-5D-5L is a validated, generalised, health related quality of life questionnaire consisting of 5 domains related to daily activities with a 5-level answer possibility, which will be converted into multi-attributed utility scores using established algorithms. The EQ-5D-5L will be scored in line with current NICE guidance, which currently stipulate that the response will be mapped to EQ-5D-3L utilities using the Hernandez Alava method (e.g. via eq5dmap in Stata and eq5d in the R software) (4).
- Pain and irritation VAS score: A VAS score used to measure patient reported knee pain and irritation. Patients will be requested to mark on a scale of 0 (no pain) to 10 (worst possible pain).
- UCLA Activity Score: The UCLA Activity score is a scale ranging from 1 to 10. The patient indicates their most appropriate activity level, with 1 defined as “wholly inactive, dependent on others, and cannot leave residence” and 10 defined as “regularly participates in impact sports”. Treat as categorical.
- Satisfaction and Transition questionnaire: To measure patient satisfaction and transition, participants will be asked how satisfied they were with the treatment they received and based on their experience, how willing they would be to have the operation again.

- `o` This questionnaire consists of 5 questions, all in a likert scale with between 3 and 6 answers. Responses to each question will be presented separately:

How satisfied are you with your knee after your HTO surgery?

How are the problems related to your knee now, compared with before your knee surgery?

If you could go back in time, would you still choose to have the knee operation?

In general, would you say that your health is:

Compared to one year ago, how would you rate your health in general now?

- Health Resource Use: No formal health economic evaluation will be performed in the remit of this trial. This study will collect information on participants’ health resource use, including time in operating theatre, visits to primary care, and hospital care services, during the follow-up. Summaries will be presented by trial arm and mean differences with 95% confidence intervals.
- Safety: Safety data will be collected throughout the duration of the trial, to determine the rates of Adverse Events and Adverse Device Effects.

### 6.2 Analysis Methods

#### 6.2.1 Assessments of normality

Given the sample size and the methods being used no formal checks of assumptions will be carried out. As the sample size is small, a non-parametric methods (Mann-Whitney), will be used and medians and IQRs will be also reported for each treatment allocation (in addition to mean and SD) as a sensitivity analysis.

#### 6.2.2 Primary Outcome - The absolute difference between achieved and planned correction at nine months post-randomisation (six months post-surgery)

Unadjusted summary statistics will be displayed by treatment allocation using means, standard deviations, and IQR. Linear regression model will be used to compare the treatment groups including all patients. The model will be adjusted for baseline misalignment only. Results from this model will be presented as adjusted mean difference in the absolute difference between achieved and planned correction (termed as “misalignment” from target value) between the groups; with corresponding 95% confidence intervals and p-values.

As a supporting analysis, a Mann-Whitney test comparing the primary outcome at 9 months will also be presented. These analyses will be performed on the ITT population, and no imputation of missing data is planned.

The primary analysis of absolute misalignment will be measured separately by two blinded reviewers, an Altman-Bland plot will be used to assess the agreement and discrepancies.

#### 6.2.3 Secondary outcomes

The key secondary outcome, i.e. number of patients (%) achieving a pre-specified difference between planned and achieved coronal plane correction at nine months post-randomisation, will be analysed by logistic regression model. The model will be adjusted for the randomisation factors (sex, age, and BMI). The results of this analysis will be presented as adjusted odds ratio, with corresponding 95% confidence intervals and p-values. The unadjusted risk differences will also be provided to ensure that both relative and absolute effect sizes are presented.

Continuous clinical secondary outcomes (change in coronal plane location of peak loading, range of motion, calibration) will be analysed using a linear regression model to compare treatment groups, adjusting for the randomisation factors (sex, age, and BMI) and baseline misalignment.

Continuous patient reported outcomes (KOOS sub-scale and summary scores, EQ-5D-5L utility score, EQ-5D-5L VAS, pain and irritation VAS) will be analysed using a linear regression model. The model will include fixed effects to adjust for sex, age, BMI, and baseline misalignment. The adjusted mean differences will be presented with 95% confidence intervals and p-values.

The UCLA Activity Score and the satisfaction and transition questionnaire will be analysed using a linear regression model, adjusting for sex, age, BMI, and baseline misalignment. Adjusted odds ratios, 95% confidence intervals and p-values will be presented.

#### 6.2.4 Safety outcomes

Intra-operative complications, device related complication, general AEs and SAEs will be presented as the numbers and percentages of participants who experienced at least one. Safety data will be analysed using a logistic regression model adjusting for age, sex, BMI and baseline misalignment, and will be presented as adjusted odds ratios with 95% confidence intervals and p-values. The unadjusted risk differences will also be provided.

### 6.3 Missing Data

The number and percentage of individuals with missing data for each outcome at each time point will be summarised by intervention. Missing data will be assumed to be missing at random, but the nature of this assumption will be explored, and if judged appropriate, missing data for the primary outcome and any key secondary outcomes will be imputed under a variety of different missing data assumptions as part of sensitivity analysis.

### 6.4 Sensitivity Analysis

The primary outcome analysis will be redone with the addition of the randomisation factors. The model will adjust for baseline misalignment, sex, age, and BMI.

The primary outcome (absolute difference) analysis, and key secondary outcome (% participants achieving pre-specified difference) analysis will be repeated for the per-protocol population. Sensitivity analysis will be undertaken for the primary and key secondary outcome by imputing data under different simple imputation missing not-at-random assumptions.

### 6.5 Pre-specified Subgroup Analysis

There are no sub-group analyses planned for this study.

### 6.6 Supplementary/Additional Analyses and Outcomes

There are no planned supplementary analyses for this study.

### 6.7 Health Economics and Cost Effectiveness (where applicable)

Health resource use data will be presented with numbers and percentages. This will cover operation times, inpatient visits and outpatient visits during the follow-up period. The number of GP clinic visits, home visits and telephone consultations will also be recorded alongside the number of practice nurse visits, practice nurse consultations, district nurse visits, physiotherapy visits, occupational therapy visits and counsellor/psychiatrist/psychologist visits.

Device deficiencies will also be descriptively summarised. This includes deficiencies caused by identity, quality, durability, reliability, safety, performance, malfunction, user error and inadequate labelling.

## 7. Validation of the Primary analysis

To validate the primary outcome (absolute difference between planned and achieved correction between pre-op and post-op alignment) and key secondary outcomes (number (%) achieving a pre-specified difference between planned and achieved correction) a statistician not involved in the trial will independently repeat the analyses detailed in this SAP, by using different statistical software (if possible). The results will be compared and any discrepancies will be reported in the Statistical report (See OCTRU SOP STATS-005 Statistical Report).

## 8. Specification of Statistical Packages

All analysis will be carried out using appropriate validated statistical software such as STATA or R. The relevant package and version number will be recorded in the Statistical report.

## Appendix: glossary of abbreviations

SAP: Statistical Analysis Plan
DSMC: Data and Safety Monitoring Committee
TSC: Trial Steering Committee
CI: Chief Investigator
HTO: High Tibial Osteotomy
TOKA: Tailored Osteotomy Knee Alignment
ICE: Intercurrent Event
KOOS: Knee injury and Osteoarthritis Outcome Score
PROM: Patient Reported Outcome Measure
ROM: Range of Motion
UCLA: Activity Score University of Carolina, Los Angeles Activity Score
VVA: Video Vector Analysis
HKA: Hip Knee Ankle
ITT: Intention-to-Treat
PP: Per Protocol

